# Genome-wide polygenic risk score for muscle strength predicts lower risk for common diseases and longer life span among the Finnish population: a prospective population based cohort study of the 342 443 FinnGen participants

**DOI:** 10.1101/2023.03.29.23287905

**Authors:** Päivi Herranen, Kaisa Koivunen, Teemu Palviainen, FinnGen, Urho M Kujala, Samuli Ripatti, Jaakko Kaprio, Elina Sillanpää

**Author notes:** **Corresponding author:** Elina Sillanpää, Associate Professor, Faculty of Sport and Health Sciences, P.O. Box 35 (VIV), FIN-40014 University of Jyväskylä, Finland.

## Abstract

**Purpose:** To use a genome-wide polygenic risk score for hand grip strength (PRS HGS) to investigate whether the muscle strength genotype predicts the most common age related noncommunicable diseases, survival from acute adverse health events, and all cause mortality.

**Methods:** This study consisted of 342 443 Finnish biobank participants from FinnGen Data Freeze 10 (53% women) aged 40 to 108 with combined genotype and health registry data. Associations were explored with a linear or Cox proportional hazards regression models.

**Results:** A higher PRS HGS predicted a lower body mass index (BMI) (β = −0.112 kg/m^2^, standard error (SE) = 0.017, *P* = 1.69 × 10^−11^) in women but not in men (β = 0.004 kg/m^2^, *P* = 0.768, sex by PRS HGS interaction: *P* = 2.12 × 10^−07^). In all participants, a higher PRS HGS was associated with a lower risk for obesity diagnosis (hazard ratio 0.94, 95% confidence interval 0.93 to 0.95), type 2 diabetes (0.95, 0.94 to 0.96), ischemic heart diseases (0.97, 0.96 to 0.97), hypertension (0.97, 0.96 to 0.97), stroke (0.97, 0.96 to 0.98), asthma (0.94, 0.93 to 0.95), chronic obstructive pulmonary disease (0.94, 0.92 to 0.95), polyarthrosis (0.90, 0.88 to 0.92), knee arthrosis (0.98, 0.97 to 0.99), rheumatoid arthritis (0.95, 0.94 to 0.97), osteoporosis (0.95, 0.93 to 0.97), falls (0.98, 0.98 to 0.99), depression (0.95, 0.94 to 0.96), and vascular dementia (0.93, 0.89 to 0.96). In women only, a higher PRS predicted a lower hazard for any dementia (0.94, 0.92 to 0.96) and Alzheimer’s disease (0.96, 0.93 to 0.98). Participants with a higher PRS HGS had a decreased risk of cardiovascular (0.96, 0.95 to 0.98) and all cause mortality (0.97, 0.96 to 0.98). However, the predictive value of the PRS HGS for mortality was not pronounced after adverse acute health events compared to the non-diseased period.

**Conclusions:** The genotype that supports higher muscle strength protects against many future health adversities. Further research is needed to investigate whether or how a favourable lifestyle modifies this intrinsic capacity to resist diseases, and if the impacts of lifestyle behaviour on health differ due to polygenic risk.

## INTRODUCTION

Noncommunicable diseases, which are defined as medical conditions with long durations, are the leading cause of death and disability worldwide. World Health Organization further specifies that noncommunicable diseases are the result of a combination of genetic, physiological, environmental and behavioural factors [1]. Among such factors is voluntary maximal muscle strength. It is the highest force a person can exhibit. Muscle strength may reflect the individual’s intrinsic physiological capacity to resist functional decline into critical disease and disability levels, but also to recover from episodes of poor health over the lifespan [2,3]. In particular, low hand grip strength (HGS), measured at any time during adulthood, predicts future adversities and premature mortality [4–6]. HGS correlates strongly with total muscle strength and is widely used as an indicator of general health and vigour [4]. Reduced HGS has been found to be associated with a higher occurrence of cardiovascular diseases [7–11], diabetes [12,13], pulmonary diseases [8], cancers [14] as well as depression and anxiety disorders [15], and dementia [16]. Higher HGS has been indicated to be associated with a lower risk for all cause mortality, especially cardiovascular deaths, as well as death due to pulmonary diseases and cancer [8,9,11]. HGS has also been shown to predict falls [17] and fracture risk [18], and higher HGS assessed before a bone fracture has been observed to be associated with enhanced survival during recovery [19]. Furthermore, low HGS is associated with an increased risk of being hospitalised [20] and a longer time to discharge [21].

Both the HGS and its trajectory over the life course are highly individual and are affected by genes, accumulated lifestyle exposures, the burden of diseases, and progressive physiological ageing changes [22,23]. Hence, HGS is a multifactorial and a polygenic trait. It has a substantial genetic component (heritability estimates of [h^2^] 30–65%) according to twin studies [24]. No known genetic variants of large effect have been found [25], but genome-wide association studies (GWASs) of HGS have identified a large number of common variants each of a small effect [26,27]. Thus, polygenic risk scores (PRSs) can summarise an individual’s genetic risk for a trait into a single value estimate [28]. Recently, we constructed a PRS for HGS and showed that it also predicts variation in measures of muscle strength other than HGS and is associated with better physical functioning, as well as a lower risk of functional limitations among older women [29]. This suggests that the PRS HGS may be used as an estimate of the muscle strength genotype. Individual PRSs can also be used to study genetic pleiotropy, that is, whether the same genetic variation overlaps in two or more traits [30]. HGS may share a common genetic base with several disease and disability outcomes and subsequent mortality [26,27].

Despite considerable progress in muscle strength research, the genetic aspects of muscle strength are not yet fully understood and might play an important role in healthy ageing. We hypothesised that genetically determined muscle strength is an important predictor of future health and lifespan. In this study, we investigated whether PRS HGS predicts common noncommunicable diseases and conditions, and mortality among the Finnish population. Furthermore, the important role of muscle strength in recovering and survival with acute diseases and conditions [3,19] suggests that individuals with higher inherited muscle strength might have a lower mortality risk after acute adverse health events. To test this hypothesis, we assessed whether the potential association between PRS HGS and mortality risk was pronounced after acute adverse health events compared to the non-diseased period.

## METHODS

### Study sample and endpoints

The data comprised 429 200 genotyped Finnish citizens from the latest data freeze of the Finnish FinnGen study (data freeze 10, autumn 2022) [31]. Genetic principal components (PCs) to correct potential confounding due to population structure [32] were available for 412 181 participants. For this study, we excluded individuals who were under 40 years old at the time of death or at the end of follow-up. The final number of participants included in this analysis was 342 443 individuals.

FinnGen includes prospective epidemiological cohorts, disease-based cohorts, and hospital biobank samples (supplemental file 1). In the FinnGen study, genome information is linked by a unique national personal identification number with national hospital discharge (from 1968), causes of death (from 1969), and cancer (from 1953) registers, and the Social Insurance Institute of Finland (Kela) medication reimbursement (from 1965) and prescribed medicine purchase (from 1995) registers.

Endpoint definitions were based on the *International Statistical Classification of Diseases and Related Health Problems* (ICD-8, ICD-9, and ICD-10) codes. In this study, selected endpoints for the analysis were based on the leading causes of death [33,34] and on the noncommunicable diseases and conditions that are considered major public health issues in Finland [35]. Endpoint definitions for selected diseases, created by panels of clinical specialists and researchers, are described in the supplemental file 2. Detailed descriptions of the ICD codes included in each endpoint can be viewed on the FinnGen website (https://www.finngen.fi/en/researchers/clinical-endpoints, -DF10). In analysis, smoking was categorized into current, former and never smokers based on self-reports.

Most of FinnGen participants have been recruited from hospital biobanks or diseased based cohorts [36], which may lead to an overestimation of absolute disease risk. To check the robustness of our primary analysis, we also conducted sensitivity analysis using a population based subset of FinnGen, the prospective epidemiological FINRISK study with 28 543 individuals. FINRISK surveys were performed in 1992, 1997, 2002, 2007, and 2012 comprised random samples of adults within five geographical areas in Finland. Additional details on the study protocol have been described in elsewhere [37].

### Genotyping, quality control, and imputation

The FinnGen individuals were genotyped with Illumina and Affymetrix chip arrays (Illumina Inc., San Diego, and Thermo Fisher Scientific, Santa Clara, CA, USA). For detailed information on genotyping, quality control, and imputation, please see supplemental file 3 and the FinnGen website (https://finngen.gitbook.io/documentation/).

### Polygenic score for HGS

We adapted a recently developed PRS for maximum HGS [29] to the FinnGen cohort. Briefly, we obtained polygenic scoring by Bayesian methodology (SBayesR) [28] using freely available GWAS summary statistics from 40-to 69-year-old participants of the Pan-UK Biobank (https://pan.ukbb.broadinstitute.org/). The data were restricted to 418 827 European individuals. The method utilises a sparse linkage disequilibrium (LD) reference panel generated by SBayesR authors. The reference panel is based on a random sample of 50 000 UK Biobank (UKBB) [38] individuals. The original summary statistics included 34 263 104 genetic variants. For computational reasons, we restricted the LD reference panel, summary statistics, and FinnGen target study samples to 1 006 473 HapMap3 [39] variants, which represent the whole genome and are well imputed for samples of European ancestry. A detailed description of the PRS HGS calculation was presented in our previous study [29].

### Statistical analyses

#### Association and survival analyses

We analysed the association between PRS HGS and BMI with linear regression models. We used Cox proportional hazards models to investigate the association between PRS HGS and disease endpoints and mortality. We assessed the proportional hazard assumptions visually by Kaplan-Meier survival curves and by Schoenfeld residuals to ensure they were not violated. We conducted all survival analyses with age as the time scale, and adjusted models for sex, collection year, the first ten genetic PCs of ancestry, and genotyping batch. Since genetic information and sex remain constant throughout the follow-up of an individual, we set the start of follow-up at birth, and it ended at the first record of the selected endpoint, death, or on 31 December 2021. To identify the possible immortal bias [40] in survival analysis, we also performed sensitivity analysis by setting age at the blood sampling for DNA analysis as the start of follow-up. We investigated potential sex differences in the effect of PRS HGS on outcome by fitting the interaction term between PRS HGS and sex into the models. We have presented the results separately for women and men only if we found a significant interaction between the PRS HGS and sex; otherwise, the results are presented adjusted for sex.

#### Time-dependent survival analysis

To analyse mortality risk before and after onset an acute disease, we used Cox regression analysis with an extension of the illness–death model [41]. We restricted acute adverse health events to ischemic heart diseases, stroke, and femur fracture, as mortality is known to increase during the first year after the event for all these diagnoses [42–44]. In the illness– death models, we set the follow-up from 40 years of age because consequences after acute adverse health events are known to be less fatal in the younger population [45]. We modelled the disease state as a time-dependent variable in a relative risk model based on a counting process formulation. The possible diseased states for the study participants are shown in figure 1. All participants started in the non-diseased state until an adverse health event occurred, or until death or end of follow-up if they did not have the event of interest. The main effects of PRS HGS indicate mortality risk as PRS changes, and the main effects of diseased states indicate mortality risk compared to non-diseased states. We used interaction terms between PRS HGS and diseased states to investigate whether the association between PRS HGS and mortality risk was different during the first post acute event year or after the first post acute event year compared to the non-diseased state.

**Figure 1.**
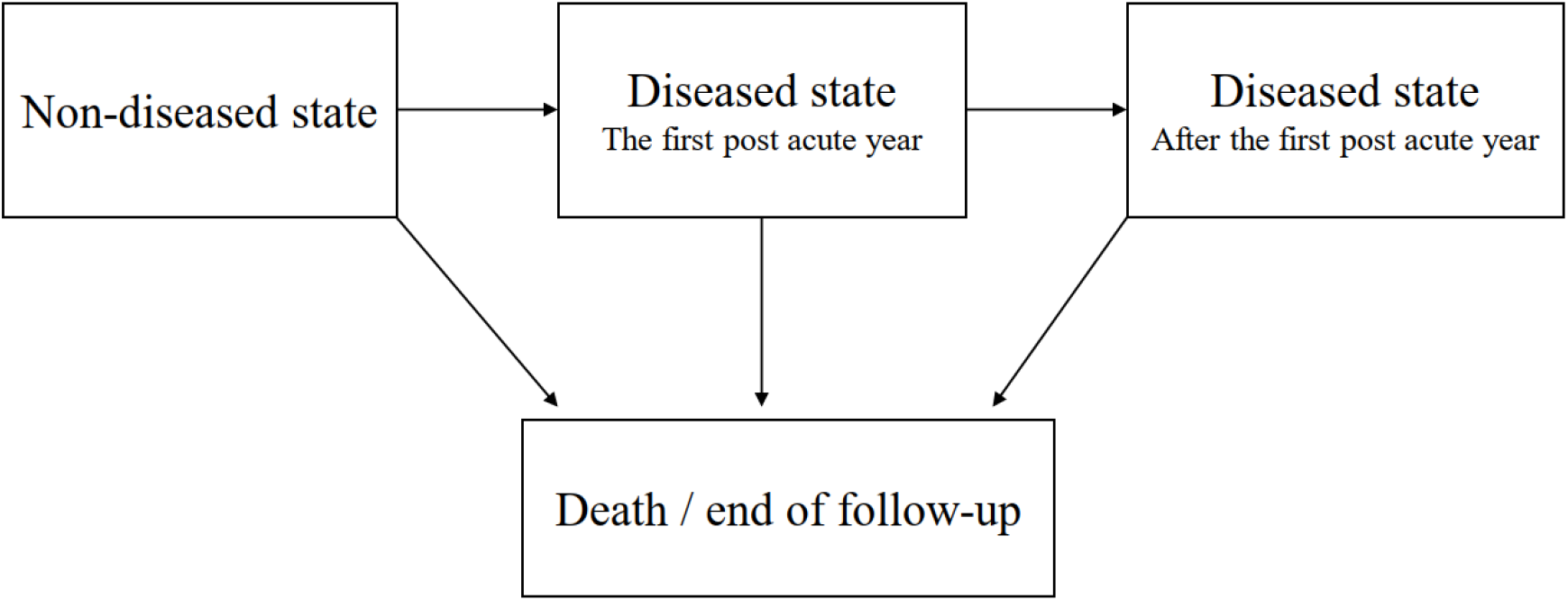
An extension of the illness–death model used in the analysis.

In all analyses, we calculated an increase in risk per 1 standard deviation (SD) change in the PRS HGS and set the significance threshold to P < 0.05. In the survival analysis, we reported the results as hazard ratios (HRs) together with 95% confidence intervals (CIs). We performed statistical analyses using R 3.6.3 with the R package survival and survminer. For the PRS calculation, we used PLINK 2.0 software.

## RESULTS

Table 1 summarises the main characteristics of the FinnGen participants. A slight majority (53.2%) were women, and the mean age at the time of death or at the end of follow-up was 66.3 years (range, 40–107.7). Of those with a known smoking status, 47.6% were never smokers. A higher PRS HGS predicted a lower risk for smoking (hazard ratios for former vs. never smokers 0.99, 0.98 to 0.99 and for current vs. never smokers 0.97, 0.97 to 0.98).

**Table 1.** Characteristics of participants in the FinnGen study.

| **Characteristics** | **All**<br>(N=342 443) | **N** | **Women**<br>(N=182 309) | **N** | **Men**<br>(N=160 134) | **N** |
| --- | --- | --- | --- | --- | --- | --- |
| Mean (SD) age (y) | 66.29 (12.90) | 342 443 | 64.72 (13.08) | 182 309 | 68.08 (12.47) | 160 134 |
| Mean (SD) BMI (kg/m<sup>2</sup>) | 27.67 (5.30) | 247 051 | 27.73 (5.86) | 123 892 | 27.62 (4.68) | 123 159 |
| Mean (SD) height (cm) | 170.3 (9.14) | 248 515 | 164 (6.31) | 124 803 | 176.7 (6.84) | 123 737 |
| Mean (SD) weight (kg) | 80.49 (17.39) | 252 839 | 74.63 (16.46) | 127 189 | 86.42 (16.27) | 125 650 |
| Smoking status N (%) |  | 202 771 |  | 100 168 |  | 102 603 |
| Never | 96 553 (47.6) |  | 60 646 (60.5) |  | 35 907 (35.0) |  |
| Former | 47 149 (23.3) |  | 20 881 (20.8) |  | 26 268 (25.6) |  |
| Current | 59 069 (29.1) |  | 18 641 (18.6) |  | 40 428 (39.4) |  |

Age at the time of death or at the end of follow-up on 31 December 2021. Phenotype data were obtained from the biobanks (https://www.finnngen.fi/en/data_protection/data-protection-statement), SD=standard deviation, BMI=body mass index.

### PRS HGS and risk for cardiometabolic and pulmonary diseases

A significant interaction effect between sex and PRS HGS was seen on BMI (*P* = 2.12 × 10^−07^). High PRS HGS predicted a lower BMI (*β* = −0.112 kg·m^−2^, SE = 0.017, *P* = 1.69 × 10^−11^, *N* = 123 892) in women, but not in men (*β* = 0.004, SE = 0.013, *P* = 0.768, *N* = 123 159). In all, a higher PRS HGS was associated with a decreased risk for obesity diagnosis (0.94, 0.93 to 0.95) and developing type 2 diabetes (0.95, 0.94 to 0.96). A higher PRS HGS predicted a systemically smaller risk for ischemic heart diseases (0.97, 0.96 to 0.97), hypertension (0.97, 0.96 to 0.97), and stroke (0.97, 0.96 to 0.98), as well as asthma (0.94, 0.93 to 0.95) and chronic obstructive pulmonary disease (COPD) (0.94, 0.92 to 0.95) (figure 2).

**Figure 2.**
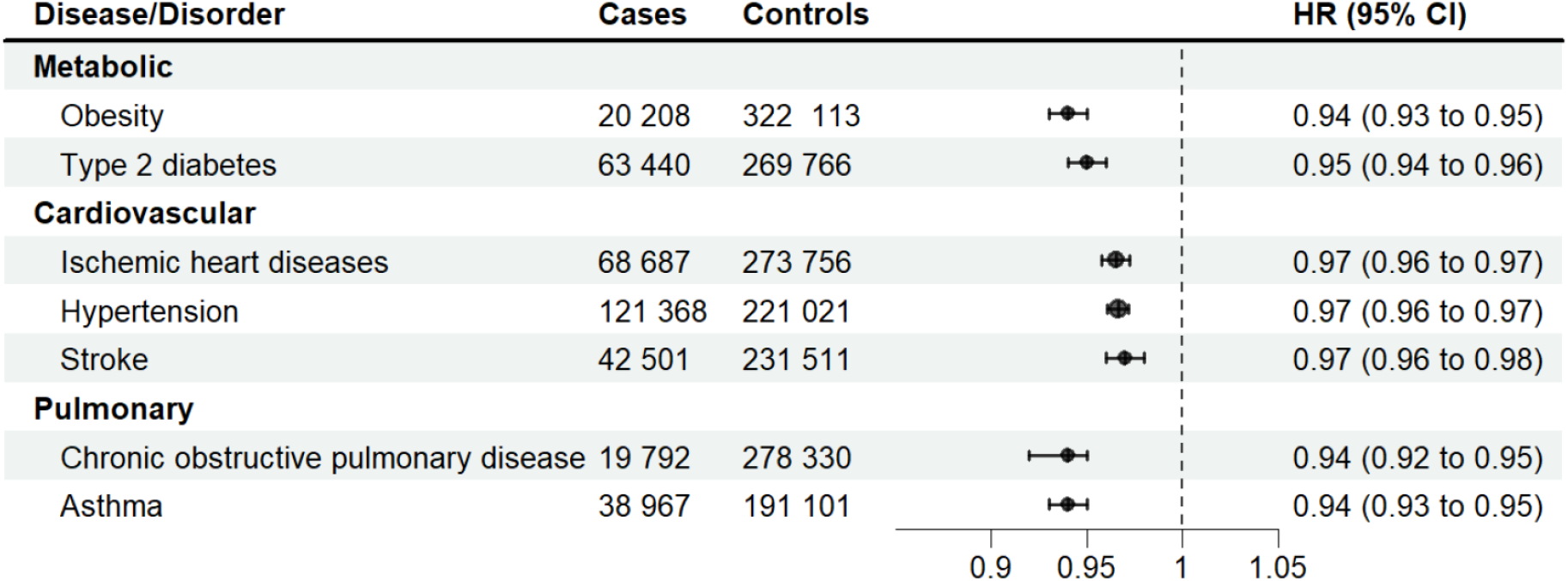
PRS HGS as a predictor of cardiometabolic and pulmonary diseases. Multivariable Cox regression analysis. Adjusted for sex, collection year, genotyping batch, and ten principal genetic components of ancestry. HR=hazard ratio, CI=confidence interval.

### PRS HGS and risk for musculoskeletal and connective tissue diseases, falls, and fractures

In all, a higher PRS HGS was associated with a lower risk for polyarthrosis (0.90, 0.88 to 0.92), knee arthrosis (0.98, 0.97 to 0.99), rheumatoid arthritis (0.95, 0.94 to 0.97), and osteoporosis (0.95, 0.93 to 0.97), but PRS HGS was not associated with a risk of hip arthrosis (1.00, 0.98 to 1.01) (figure 3).

**Figure 3.**
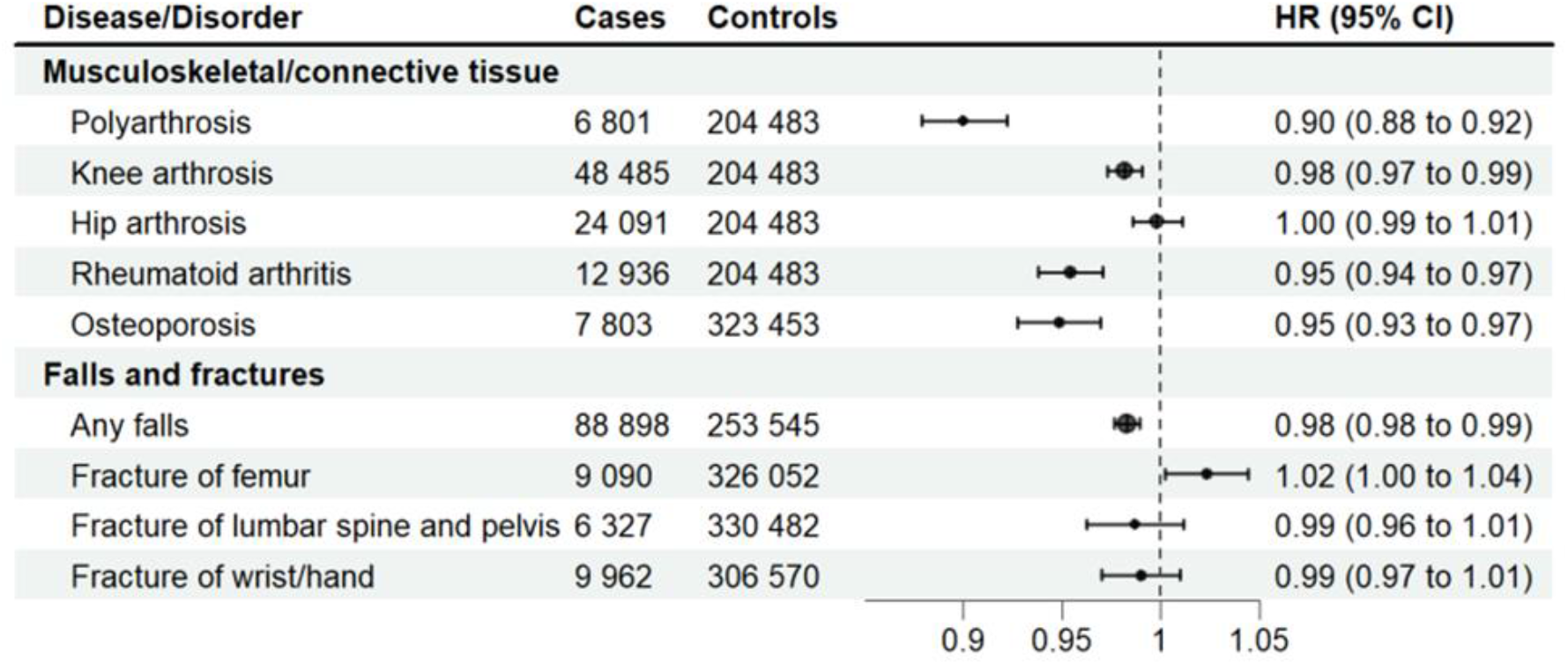
PRS HGS as a predictor of musculoskeletal and connective tissue diseases, falls, and fractures. Multivariable Cox regression analysis. Adjusted for sex, collection year, genotyping batch, and ten principal genetic components of ancestry. HR=hazard ratio, CI=confidence interval.

Genetically stronger individuals were also less likely to fall (0.98, 0.98 to 0.99), but we found no significant associations between inherited muscle strength and fractures.

### PRS HGS and risk for mental and cognitive disorders

In all, a higher PRS HGS predicted a lower risk of depression (0.95, 0.94 to 0.96) and vascular dementia (0.93, 0.89 to 0.96). Also, women with a higher genetic muscle strength genotype had a decreased risk for any dementia (0.94, 0.92 to 0.96) and Alzheimer’s disease (0.96, 0.93 to 0.98) (figure 4).

**Figure 4.**
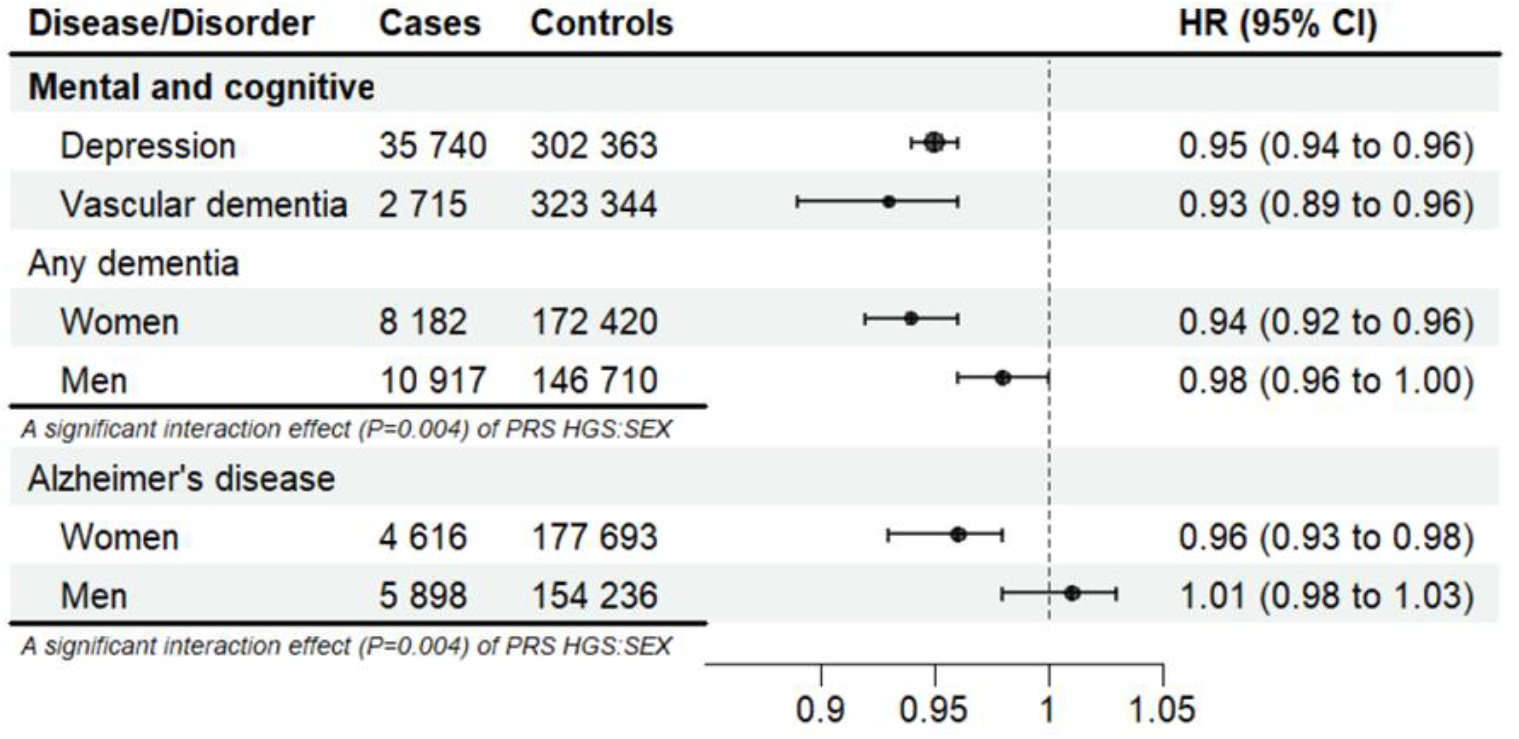
PRS HGS as a predictor of mental and cognitive disorders. Multivariable Cox regression analysis. Adjusted for sex, collection year, genotyping batch, and ten principal genetic components of ancestry. Results with a significant interaction effect (*P* <0.05) between PRS HGS and sex are presented stratified by sex. HR=hazard ratio, CI=confidence interval.

### PRS HGS and the risk of cancer and mortality

PRS HGS was not associated with the five most common cancers (figure 5).

**Figure 5.**
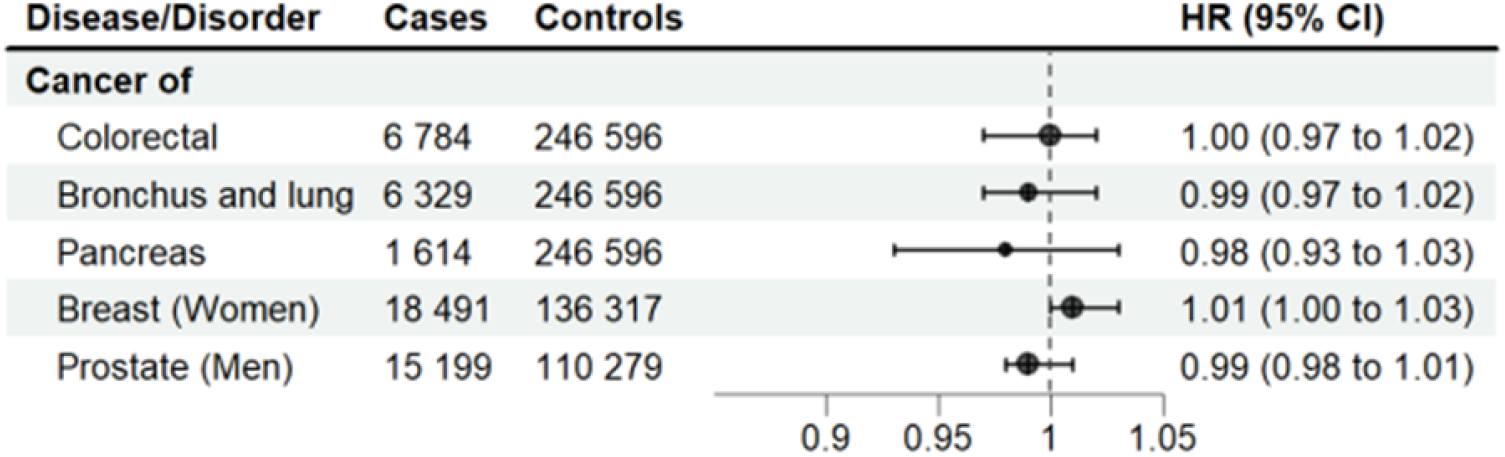
PRS HGS as a predictor of common cancers. Multivariable Cox regression analysis. Adjusted for sex, collection year, genotyping batch, and ten principal genetic components of ancestry. HR=hazard ratio, CI=confidence interval.

A higher PRS HGS was associated with a decreased risk of death due to cardiovascular causes (0.96, 0.95 to 0.98) and predicted lower all cause mortality (0.97, 0.96 to 0.98) (figure 6).

**Figure 6.**
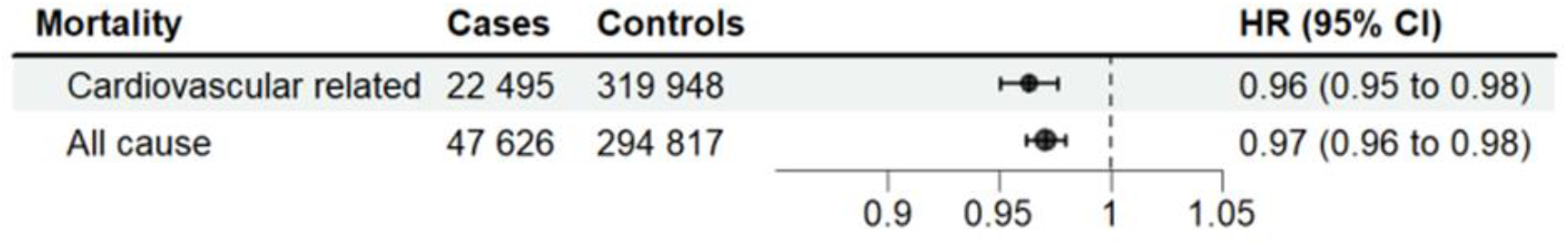
PRS HGS as a predictor of mortality. Multivariable Cox regression analysis. Adjusted for sex, collection year, genotyping batch, and ten principal genetic components of ancestry. HR=hazard ratio, CI=confidence interval.

### Mortality risk during and after the first post acute event year compared to the non-diseased period

The investigated acute adverse health events included ischemic heart disease, stroke, and femur fracture. Supplemental file 4; supplemental table 1 provides the characteristics of the participants according to diseased state (non-diseased, survived the first post acute event year, and died during the first post acute event year). In total, 9 992 (14.8%) participants who faced acute ischemic heart disease and 5 877 (14.4%) participants with stroke died during the first year after the event. Of the participants with femur fracture, 2 007 (30.8%) died during the first year post-fracture. In all participants, those who died during the first year after an acute event were significantly older and were likely to be former or current smokers compared to the first-year post acute event survivors and non-diseased participants (supplemental file 4; supplemental table 1). The association between PRS HGS and mortality was not pronounced during or after the first post acute year after the acute events compared to the non-diseased period (supplemental file 4; supplemental table 2). The predictive value of PRS HGS on mortality decreased after the first post-stroke year compared to the non-diseased period (supplemental file 4; supplemental tables 1 and 2).

### Sensitivity analysis

Supplemental file 4 contains results from the sensitivity analysis. Marked differences in results were not observed between the analysis conducted from birth and from blood sampling age or using population based FINRISK cohort. However, wide confidence intervals in sensitivity analysis indicated that prognostic imbalance with small sample size could be substantial (supplemental file 4; supplemental figures 1-15).

## DISCUSSION

We utilised a novel genome-wide polygenic scoring methodology and showed that individuals with a genotype supporting greater muscle strength have a significantly reduced risk of several age related noncommunicable diseases. Further, this genotype was associated with a lower risk of mortality due to cardiovascular causes and a lower risk of all cause mortality. We also investigated the potential role of the muscle strength genotype during recovery periods and found that higher inherited muscle strength did not predict better survival after acute adverse health events. Our results suggest that genetically inherited muscle strength may reflect an individual’s intrinsic capacity to resist pathological changes that occur over ageing, but might not reflect physical resilience, that is, the ability to recover after severe adversity.

Associations between maximal HGS and the occurrence of several noncommunicable conditions, especially cardiometabolic and pulmonary diseases, are well recognised [7–10,12,13]. The mechanisms underlying the associations between cardiorespiratory fitness and maximal muscle strength and metabolic risk factors remain unclear but are suggested to be associated with skeletal muscle metabolism, body fat content, and overall metabolic processes [46]. Our results suggest that these associations are partly explained by the genetic inheritance of muscle strength. While a family-based study by Yudkovicz et al. [10] did not find a genetic correlation between HGS and cardiovascular disease risk factors, large GWASs have recently succeeded in indicating a partly shared genetic aetiology underlying both HGS and common cardiometabolic conditions [26,27]. Lakshman Kumar et al. [47] also found that the genetic variation associated with COPD-related weight loss plays an important role in skeletal muscle regeneration and tissue remodelling. Along with the liver and kidneys, skeletal muscle has a unique ability to store glucose in the form of glycogen, making it the largest metabolic organ and important in maintaining normal blood glucose level [3]. The essential role of skeletal muscle in regulating metabolic homeostasis and respiratory mechanics may contribute to our findings and why greater muscle genotypes protect against cardiometabolic and pulmonary diseases. Our results also advance understanding regarding the partly shared genetic architecture of muscle strength and common cardiopulmonary diseases and highlight the importance of maintaining adequate muscle strength throughout the lifespan.

We found that a genotype that supports higher muscle strength predicted a lower risk of Alzheimer’s disease and dementia in women. These results expand the findings of the latest studies, which have suggested that HGS is associated with early stage cognitive dysfunctions and all-cause dementia independent of the most important sociodemographic, health, and behavioural confounders [16,48]. Furthermore, Tikkanen et al. [26] showed that HGS genetic score used in their study was significantly associated with cognitive performance, and we recently reported that PRS HGS predicts cognitive tasks in laboratory settings [29]. Neuromuscular function underlies maximal muscle strength. Thus, the connection between muscle strength and cognitive performance and disorders might be explained by neurodegenerative and neurochemical changes that affect both phenotypes [49] and/or by shared genetic variations. GWASs have indicated several loci and genes overlap highly with HGS and neuro-developmental disorders or brain function and enrichment of gene expression of brain-related transcripts [26,27]. The identified mechanisms regulate neuronal maintenance and signal transduction or are related to monogenic syndromes with the involvement of psychomotor impairment [27]. In our study, the muscle strength genotype also predicted a lower risk of depression in both sexes. This result is in line with a large study among UK Biobank participants, which showed that a higher HGS was associated with a lower incidence of depression [15]. GWASs have shown genes and gene pathways associated with synaptic structure and neurotransmission but also significant enrichment in the central nervous system and skeletal muscle tissue for variants contributing to the heritability of depressive disorders [50]. Based on our results, muscle strength, cognition functions, and depressive disorders may be partly regulated by the same genetic background.

Women and men differ in disease prevalence, manifestation, progression response to treatment, and mortality [51]. These disparities might be explained by biological sex and other genetic factors, hormonal factors, and differences in physiological characteristics, as well as gender differences in health behaviour and socio-cultural constructions during the life course [51,52]. However, the prognostic value of grip strength does not seem to differ between men and women [8,9,13]. In the current study, sex differences in the predictive ability of the muscle strength genotype were seen only for cognition disorders and BMI. BMI measures both fat-mass and fat-free mass, including muscle, and these proportions differ by sex at any given BMI value. Based on the literature, association between HGS and BMI in both genders and all age is controversial [53,54]. GWASs have suggested genetic correlation between HGS and body composition measures such as BMI, lean body mass, body fat, and waist and hip circumference [26,27], but further investigation into the association of muscle strength genotypes and body composition is needed. At the genetic level, recent studies have found small differences in genetic architecture between the sexes in a large number of human traits and diseases, but overall, genetics in most of the characteristics seem to be common between sexes [55,56].

In a broad sense, physical resilience has been determined as an individual’s ability to resist and recover from harmful health effects and maintain well-being during or after health adversities [2,57]. We hypothesised that genetically inherited muscle strength might be an indicator of physiologic reserve, which describes a biological capacity to respond to alterations in physiologic demands [2]. Based on our results, greater genetic muscle strength did not enhance survival after acute adversities but reduced the risk of developing further diseases and conditions. This may indicate that greater genetic muscle strength reflects more physical robustness, that is, one’s intrinsic ability to resist and shelter oneself from adverse health events, than physical resilience, which is considered the ability to recover or completely bounce back after health adversities [57]. Although both decline with age, it has been hypothesised that the mechanisms underlying changes in physical robustness and resilience during ageing may not be entirely the same [58].

The PRS HGS used in this study is a reliable variable that represents genetically inherited overall muscle strength [29]. It was derived from GWASs by Pan-UKBB, which were restricted to European ancestry. The Finnish population is known to be a genetic isolate with recent bottlenecks, and the frequency of less common and rare variants differs from that of other Europeans [59]. However, the rates of common variants are highly comparable to those of other European populations. It must be noted that, due to UKBB participants being volunteers, they are healthier and may be stronger compared to the general British population and individuals of the same age [60]. This suggests that the reported associations in this study may underestimate the true associations. On the other hand, a healthier base population reduces the likelihood that PRS HGS includes genetic variants that are primarily predictors of chronic diseases. In the present study, we used a study sample of over 340 000 Finnish individuals over the age of 40 and validated registry-based healthcare data [61]. A minor limitation is that we used existing FinnGen endpoints and did not exclude, for example, violent and accidental deaths or high-energy fractures from our analysis. Our results are well generalisable to Finns and probably Europeans overall because sample size covers over 11% of the same age in the entire Finnish population, and sensitivity analyses with population based FINRISK study suggest similar associations.

## Conclusion

Our results suggest that PRS HGS is an important and useful predictor of health outcomes. PRS HGS may also have potential clinical utility alongside traditional risk management in identifying high-risk individuals for common noncommunicable diseases. It could be applied in further studies to explore whether the associations between muscle strength and future health adversities are causal or are explained by shared genetic and/or environmental factors. Also, it could be used to study how lifestyle, such as physical activity, modifies human intrinsic capacity to resist diseases and whether their impact on health differs due to polygenic risk. Further research is also needed to determine whether an individual’s genetic predisposition to muscle strength affects exercise responses and trainability.

## Supporting information

FinnGen-banner_Authors

## Data Availability

Researchers can apply to use the FinnGen resource and access the data used. The Finnish biobank data can be accessed through the Fingenious® services (https://site.fingenious.fi/en/) managed by FINBB. Finnish Health register data can be applied from Findata (https://findata.fi/en/data/).

## Acknowledgements

We want to acknowledge the participants and investigators of FinnGen, FINRISK, and UK Biobank studies for their invaluable contributions to this work. The FinnGen project is funded by two grants from Business Finland (HUS 4685/31/2016 and UH 4386/31/2016) and the following industry partners: AbbVie Inc., AstraZeneca UK Ltd, Biogen MA Inc., Bristol Myers Squibb (and Celgene Corporation & Celgene International II Sàrl), Genentech Inc., Merck Sharp & Dohme LCC, Pfizer Inc., GlaxoSmithKline Intellectual Property Development Ltd., Sanofi US Services Inc., Maze Therapeutics Inc., Janssen Biotech Inc, Novartis AG, and Boehringer Ingelheim International GmbH. Following biobanks are acknowledged for delivering biobank samples to FinnGen: Auria Biobank (www.auria.fi/biopankki), THL Biobank (www.thl.fi/biobank), Helsinki Biobank (www.helsinginbiopankki.fi), Biobank Borealis of Northern Finland (https://www.ppshp.fi/Tutkimus-ja-opetus/Biopankki/Pages/Biobank-Borealis-briefly-in-English.aspx), Finnish Clinical Biobank Tampere (www.tays.fi/en-US/Research_and_development/Finnish_Clinical_Biobank_Tampere), Biobank of Eastern Finland (www.ita-suomenbiopankki.fi/en), Central Finland Biobank (www.ksshp.fi/fi-FI/Potilaalle/Biopankki), Finnish Red Cross Blood Service Biobank (www.veripalvelu.fi/verenluovutus/biopankkitoiminta), Terveystalo Biobank (www.terveystalo.com/fi/Yritystietoa/Terveystalo-Biopankki/Biopankki/) and Arctic Biobank (https://www.oulu.fi/en/university/faculties-and-units/faculty-medicine/northern-finland-birth-cohorts-and-arctic-biobank). All Finnish Biobanks are members of BBMRI.fi infrastructure (www.bbmri.fi). Finnish Biobank Cooperative -FINBB (https://finbb.fi/) is the coordinator of BBMRI-ERIC operations in Finland. The Finnish biobank data can be accessed through the Fingenious^®^ services (https://site.fingenious.fi/en/) managed by FINBB.

## Author contributions

ES and PH conceived of the idea for the study. PH, KK, and ES contributed to the design of the study and interpreted the data. PH performed the statistical analysis, assisted by KK, TP, JK, SR, and ES. PH and ES drafted the first version of the manuscript, and KK contributed significantly to the writing. All authors reviewed the manuscript and revised it critically for important intellectual content. All of the authors have also approved the conducted analyses and the final version of the manuscript to be published. PH, TP, JK, SR, and ES had full access to all the data in the study and took responsibility for the integrity of the data and the accuracy of the data analysis. All authors meet authorship criteria and that no others meeting the criteria have been omitted.

## Ethical approval

Patients and control subjects in FinnGen and FINRISK provided informed consent for biobank research, based on the Finnish Biobank Act. Alternatively, separate research cohorts, collected prior the Finnish Biobank Act came into effect (in September 2013) and start of FinnGen (August 2017), were collected based on study-specific consents and later transferred to the Finnish biobanks after approval by Fimea (Finnish Medicines Agency), the National Supervisory Authority for Welfare and Health. Recruitment protocols followed the biobank protocols approved by Fimea. The Coordinating Ethics Committee of the Hospital District of Helsinki and Uusimaa (HUS) statement number for the FinnGen study is Nr HUS/990/2017. The FinnGen study is approved by Finnish Institute for Health and Welfare (permit numbers: THL/2031/6.02.00/2017, THL/1101/5.05.00/2017, THL/341/6.02.00/2018, THL/2222/6.02.00/2018, THL/283/6.02.00/2019, THL/1721/5.05.00/2019 and THL/1524/5.05.00/2020), Digital and population data service agency (permit numbers: VRK43431/2017-3, VRK/6909/2018-3, VRK/4415/2019-3), the Social Insurance Institution (permit numbers: KELA 58/522/2017, KELA 131/522/2018, KELA 70/522/2019, KELA 98/522/2019, KELA 134/522/2019, KELA 138/522/2019, KELA 2/522/2020, KELA 16/522/2020), Findata permit numbers THL/2364/14.02/2020, THL/4055/14.06.00/2020, THL/3433/14.06.00/2020, THL/4432/14.06/2020, THL/5189/14.06/2020, THL/5894/14.06.00/2020, THL/6619/14.06.00/2020, THL/209/14.06.00/2021, THL/688/14.06.00/2021, THL/1284/14.06.00/2021, THL/1965/14.06.00/2021, THL/5546/14.02.00/2020, THL/2658/14.06.00/2021, THL/4235/14.06.00/2021, Statistics Finland (permit numbers: TK-53-1041-17 and TK/143/07.03.00/2020 (earlier TK-53-90-20) TK/1735/07.03.00/2021, TK/3112/07.03.00/2021) and Finnish Registry for Kidney Diseases permission/extract from the meeting minutes on 4^th^ July 2019. The Biobank Access Decisions for FinnGen samples and data utilized in FinnGen Data Freeze 10 include: THL Biobank BB2017_55, BB2017_111, BB2018_19, BB_2018_34, BB_2018_67, BB2018_71, BB2019_7, BB2019_8, BB2019_26, BB2020_1, BB2021_65, Finnish Red Cross Blood Service Biobank 7.12.2017, Helsinki Biobank HUS/359/2017, HUS/248/2020, HUS/150/2022 § 12, §13, §14, §15, §16, §17, §18, and §23, Auria Biobank AB17-5154 and amendment #1 (August 17 2020) and amendments BB_2021-0140, BB_2021-0156 (August 26 2021, Feb 2 2022), BB_2021-0169, BB_2021-0179, BB_2021-0161, AB20-5926 and amendment #1 (April 23 2020)and it’s modification (Sep 22 2021), Biobank Borealis of Northern Finland_2017_1013, 2021_5010, 2021_5018, 2021_5015, 2021_5023, 2021_5017, 2022_6001, Biobank of Eastern Finland 1186/2018 and amendment 22 § /2020, 53§/2021, 13§/2022, 14§/2022, 15§/2022, Finnish Clinical Biobank Tampere MH0004 and amendments (21.02.2020 & 06.10.2020), §8/2021, §9/2022, §10/2022, §12/2022, §20/2022, §21/2022, §22/2022, §23/2022, Central Finland Biobank 1-2017, and Terveystalo Biobank STB 2018001 and amendment 25^th^ Aug 2020, Finnish Hematological Registry and Clinical Biobank decision 18^th^ June 2021, Arctic biobank P0844: ARC_2021_1001.

