## Supplemental file 1 for "Genome-wide polygenic risk score for muscle strength predicts lower risk for common diseases and longer life span among the Finnish population: a prospective population based cohort study of the 342 443 FinnGen participants"

Supplemental file 1. List of FinnGen Data Freeze 10 cohorts.

ARCTIC BIOBANK NFBC1966

ARCTIC BIOBANK NFBC1986

AURIA BIOBANK

BIOBANK OF EASTERN FINLAND

BLOOD SERVICE BIOBANK

BOREALIS BIOBANK

CENTRAL FINLAND BIOBANK

HELSINKI BIOBANK

TAMPERE BIOBANK

TERVEYSTALO BIOBANK

THL BIOBANKS:

ATBC

BOTNIA

COROGENE

FINHEALTH 2017

FINHIT

FinIPF

FINRISK 1992-2012

GENERISK

HEALTH 2000/2011

HHS

KUUSAMO (=FR11)

MIGRAINE

SUPER

T1D

FINNISH TWIN COHORT
