## Supplemental file 2 for "Genome-wide polygenic risk score for muscle strength predicts lower risk for common diseases and longer life span among the Finnish population: a prospective population based cohort study of the 342 443 FinnGen participants"

Supplemental file 2. FinnGen endpoint definitions.

Below are the FinnGen endpoint definitions for selected metabolic, cardiovascular, pulmonary as well as musculoskeletal and connective tissue diseases and cancers, which were used in our analysis. Endpoints are based on the Hospital Discharge registry, the Cause of Death registry, Cancer registry and the Social Insurance Institution of Finland (KELA) registry for reimbursements of medical expenses. More detailed information about the FinnGen endpoints definitions and codes can be found in the FinnGen webpages https://risteys.finngen.fi/ .

*Metabolic diseases*

**Obesity**

- Definition: A disorder involving an excessive amount of body fat
- FinnGen code: E4_Obesity
- Hospital Discharge registry & Cause of Death registry:
    - E66

**Type 2 diabetes**

- Definition: type II diabetes mellitus: A type of diabetes mellitus that is characterized by insulin resistance or desensitization and increased blood glucose levels. This is a chronic disease that can develop gradually over the life of a patient and can be linked to both environmental factors and heredity.
- FinnGen code: T2D
- Include endpoints: T2D_WIDE, E4_DM2

*Cardiovascular diseases*

**Ischemic heart diseases**

- Definition: coronary thrombosis: Coagulation of blood in any of the coronary vessels. The presence of a blood clot (thrombus) often leads to myocardial infarction.
- FinnGen code: I9_ISCHHEART
- Include endpoints: I9_ANGINA, I9_MI, I9_MI_STRICT, I9_MI_COMPLICATIONS, I9_POSTAMI, I9_CORATHER, I9_REVASC
- Hospital Discharge registry & Cause of Death registry:
    - ICD-10: 120-125

**Hypertension**

- Definition: Persistently high systemic arterial blood pressure. Based on multiple readings (blood pressure determination), hypertension is currently defined as when systolic pressure is consistently greater than 140 mm Hg or when diastolic pressure is consistently 90 mm Hg or more.
- FinnGen code: I9_HYPTENS
- Hospital Discharge registry & Cause of Death registry:
    - ICD-10: I10-I15, I67.4
    - ICD-9: 4019X|4029A|4029B|4039A|4040A|4059A|4059B|4372A|4059X
    - ICD-8: 400|401|402|403|404

**Stroke**

- Definition: No definition available
- FinnGen code: C_STROKE

- Include endpoints: I9_SAH, I9_ICH, I9_OTHINTRACRA, I9_STR_EXH, I9_STR_SAH, I9_TIA

*Pulmonary diseases*

**COPD**

- Definition: chronic obstructive pulmonary disease: A chronic and progressive lung disorder characterized by the loss of elasticity of the bronchial tree and the air sacs, destruction of the air sacs wall, thickening of the bronchial wall, and mucous accumulation in the bronchial tree. The pathologic changes result in the disruption of the air flow in the bronchial airways. Signs and symptoms include shortness of breath, wheezing, productive cough, and chest tightness. The two main types of chronic obstructive pulmonary disease are chronic obstructive bronchitis and emphysema.
- FinnGen code: J10_COPD
- Include endpoints: J10_EMPHYSEMA, J10_COPDNAS

**Asthma**

- Definition: A bronchial disease that is characterized by chronic inflammation and narrowing of the airways, which is caused by a combination of environmental and genetic factors resulting in recurring periods of wheezing (a whistling sound while breathing), chest tightness, shortness of breath, mucus production and coughing. The symptoms appear due to a variety of triggers such as allergens, irritants, respiratory infections, weather changes, exercise, stress, reflux disease, medications, foods and emotional anxiety.
- FinnGen code: J10_ASTHMA
- Hospital Discharge registry & Cause of Death registry:
    - ICD-10: J45-J46
    - ICD-9: 493
    - ICD-8:493

*Musculoskeletal and connective tissue diseases*

**Arthrosis**

- Definition: Hemarthrosis: Bleeding into the joints. It may arise from trauma or spontaneously in patients with hemophilia.
- FinnGen code: M13_ARTHROSIS
- Include endpoints: M13_ARTHROSIS_POLY, M13_ARTHRTOSIS_COX, M13_ARTHROSIS_KNEE, M13_ARTHROSIS_OTH

**Polyarthrosis**

- Definition: No definition available
- FinnGen code: M13_ARTHROSIS_POLY
- Hospital Discharge registry & Cause of Death registry:
    - ICD-10: M15
    - ICD-9: 7151L|7152L
    - ICD-8: 71303

**Knee arthrosis**

- Definition: spondyloarthropathy: A group of inflammatory rheumatic diseases associated with arthritis and enthesitis, and often involving the axial skeleton. The most common form of spondyloarthritis is ankylosing spondylitis. Other forms include axial spondyloarthritis, peripheral spondyloarthritis, reactive arthritis, psoriatic arthritis/spondylitis and enteropathic arthritis/spondylitis.
- FinnGen code: M13_ARTHROSIS_KNEE
- Hospital Discharge registry & Cause of Death registry:

- ICD-10: M17
- ICD-9: 7151F|7152F
- ICD-8: 71301

**Hip arthrosis**

- Definition: osteoarthritis: A noninflammatory degenerative joint disease occurring chiefly in older persons, characterised by degeneration of the articular cartilage, hypertrophy of bone at the margins and changes in the synovial membrane. It is accompanied by pain and stiffness, particularly after prolonged activity.
- FinnGen code: M13_ARTHTROSIS_COX
- Hospital Discharge registry & Cause of Death registry:
    - ICD-10: M16
    - ICD-9: 7151E|7152E
    - ICD-8: 71300

**Rheumatoid arthritis**

- Definition: rheumatoid arthritis: A chronic, systemic autoimmune disorder characterized by inflammation in the synovial membranes and articular surfaces. It manifests primarily as a symmetric, erosive polyarthritis that spares the axial skeleton and is typically associated with the presence in the serum of rheumatoid factor.
- FinnGen code: M13_RHEUMA
- Hospital Discharge registry & Cause of Death registry:
    - ICD-10: M05, M06
    - ICD-9: 7140A|7140B|7241|7142
    - ICD-8: 712[1-3]

**Osteoporosis**

- Definition: A condition of reduced bone mass, with decreased cortical thickness and a decrease in the number and size of the trabeculae of cancellous bone (but normal chemical composition), resulting in increased fracture incidence. Osteoporosis is classified as primary (Type 1, postmenopausal osteoporosis; Type 2, age-associated osteoporosis; and idiopathic, which can affect juveniles, premenopausal women, and middle-aged men) and secondary osteoporosis (which results from an identifiable cause of bone mass loss).
- FinnGen code: M13_OSTEOPOROSIS
- Hospital Discharge registry & Cause of Death registry:
    - ICD-10: M80, M81, M82
    - ICD-9: 733[0-1]
    - ICD-8: 7230|72391

*Falls and fractures*

**Falls**

- Definition: Falls/tendency to fall
- FinnGen code: FALLS
- Hospital Discharge registry & Cause of Death registry:
    - ICD-10: R29, W00-W19

**Femur fracture**

- Definition: No definition available.
- FinnGen code: ST19_FRACT_FEMUR
- Hospital Discharge registry & Cause of Death registry:
    - ICD-10: S72
    - ICD-9: 820

- ICD-8: 820

**Fracture of lumbar sacral and pelvis**
- Definition: No definition available.
- FinnGen code: ST19_FRACT_LUMBAR_SPINE_PELVIS
- Hospital Discharge registry & Cause of Death registry:
    - ICD-10: S32

**Fracture at wrist and hand level**
- Definition: No definition available.
- FinnGen code: ST19_FRACT_WRIST_HAND_LEVEL
- Hospital Discharge registry & Cause of Death registry:
    - ICD-10: S62

*Mental and cognitive disorders*

**Depression**
- Definition: unipolar depression: A mood disorder having a clinical course involving one or more episodes of serious psychological depression that last two or more weeks each, do not have intervening episodes of mania or hypomania, and are characterized by a loss of interest or pleasure in almost all activities and by some or all of disturbances of appetite, sleep, or psychomotor functioning, a decrease in energy, difficulties in thinking or making decisions, loss of self-esteem or feelings of guilt, and suicidal thoughts or attempts.
- FinnGen code: F5_DEPRESSIO
- Include endpoint: F5_DEPRESSION_PSYCHOTIC
- Hospital Discharge registry & Cause of Death registry:
    - ICD-10: F32, F33
    - ICD-9: 2961|2968|3004

**Alzheimer's disease**
- Definition: A progressive, neurodegenerative disease characterized by loss of function and death of nerve cells in several areas of the brain leading to loss of cognitive function such as memory and language.
- FinnGen code: G6_ALZHEIMER
- Hospital Discharge registry & Cause of Death registry:
    - ICD-10: G30
    - ICD-9: 3310

**Dementia**
- Definition: obsolete_dementia: ['An acquired organic mental disorder with loss of intellectual abilities of sufficient severity to interfere with social or occupational functioning. The dysfunction is multifaceted and involves memory, behavior, personality, judgment, attention, spatial relations, language, abstract thought, and other executive functions. The intellectual decline is usually progressive, and initially spares the level of consciousness.']
- FinnGen code: F5_DEMENTIA
- Hospital Discharge registry & Cause of Death registry:
    - ICD-10: F00-F09
    - ICD-9: 290|3310|4378A
    - ICD-8: 290

- KELA reimbursements: Kela codes
    - 307
- Medicine purchases : ATC
    - N06D

**Vascular dementia**

- Definition: A degenerative vascular disorder affecting the brain. It is caused by the blockage of the blood supply to the brain. It is manifested with decline of memory and cognitive functions.
- FinnGen code: I9_VASCDEM
- Hospital Discharge registry & Cause of Death registry:
    - ICD-10: F01

*Cancers*

**Colorectal cancer (controls excluding all cancers)**

- Definition: No definition available.
- FinnGen code: C3_COLORECTAL_EXALLC
- Include endpoints: C3_COLON, C3_RECTOSIGMOID_JUNCTION, C3_RECTUM

**Malignant neoplasm of bronchus and lung (controls excluding all cancers)**

- Definition: No definition available.
- FinnGen code: C3_BRONCHUS_LUNG_EXALLC
- Cause of Death registry:
    - ICD-10: C34
    - ICD-9: 162
    - ICD-8: 152
    - excluded ICD-9: 1620
    - excluded ICD-8: 1620
- Cancer registry:
    - Topography ICD-O-3 :C34
    - Morphology ICD-O-3: ANY
    - Behaviour codes: 3 (Levels: 0 = “Benign”; 1 = “Semimalignant”; 2 = “Carcinoma in situ”; 3 = “Malignant”)

**Malignant neoplasm of pancreas (controls excluding all cancers)**

- Definition: No definition available.
- FinnGen code: C3_PANCREAS_EXALLC
- Cause of Death registry:
    - ICD-10: C25
    - ICD-9: 157
    - ICD-8: 157
- Cancer registry:
    - Topography ICD-O-3 :C25
    - Morphology ICD-O-3: ANY
    - Behaviour codes: 3

**Malignant neoplasm of breast (controls excluding all cancers)**

- Definition: No definition available.
- FinnGen code: C3_BREAST_EXALLC
- Cause of Death registry:
    - ICD-10: C50
    - ICD-9: 174
    - ICD-8: 174
- KELA reimbursements: KELA codes:

    - 115
- Cancer registry:
    - Topography ICD-O-3 :C50
    - Morphology ICD-O-3: ANY
    - Behaviour codes: [23]

**Malignant neoplasm of prostate (controls excluding all cancers)**

- Definition: No definition available.
- FinnGen code: C3_PROSTATE_EXALLC
- Cause of Death registry:
    - ICD-10: C61
    - ICD-9: 185
    - ICD-8: 185
- KELA reimbursements: KELA codes:
    - 116
- Cancer registry:
    - Topography ICD-O-3 :C61
    - Morphology ICD-O-3: ANY
    - Behaviour codes: 3

*Mortality*

**Death due to cardiac causes**

- Definition: No definition available.
- FinnGen code: I9_K_CARDIAC
- Cause of Death registry:
    - ICD-10: I00-I02, I05-I09, I10-I15, I20-I25, I26-I28, I30-I52, R96, R98, R99
    - ICD-9: 39|4[0-2]|79[8-9]
    - ICD-8: 39|4[0-2]|79[8-9]

**All-cause mortality**

- Definition: Any death
- FinnGen code: DEATH
- Cause of Death registry:
    - ICD-10: ANY
    - ICD-9: ANY
    - ICD-8: ANY
