## Supplemental file 3 for "Genome-wide polygenic risk score for muscle strength predicts lower risk for common diseases and longer life span among the Finnish population: a prospective population based cohort study of the 342 443 FinnGen participants"

Supplemental file 3. Genotyping and quality control of the FinnGen data.

Chip genotyping was done using several Illumina and Affymetrix FinnGen Axiom arrays. The algorithms for genotype calling were GenCall or GenCall+zCall for Illumina and AxiomGT1 for Affymetrix chip genotypes. The genome build of all genotypes were set to GRCh38/hg38. Quality controls exclusions were done sample-wise: samples with call rate below 95% and heterozygosity test method-of-moments F coefficient estimate value deviated more than ±4SD from the mean were removed along with the samples which failed sex check or were among the multi-dimensional scaling principal component analysis outliers, and in variant-wise: variants with call rate below 98%, minor allele count below 3 and Hardy-Weinberg Equilibrium p-value lower than 1e-06 were removed.

Pre-phasing was performed using Eagle v2.3.5 and imputation with Beagle v4.1 (protocol described in dx.doi.org/10.17504/protocols.io.xbgfijw) using Sisu v4 as reference panel which consist of 8,554 Finnish whole genome sequences (depth up to 30x). As post-imputation quality control, variants with imputation quality score below 0.7 were removed.
