## Supplemental file 4 for "Genome-wide polygenic risk score for muscle strength predicts lower risk for common diseases and longer life span among the Finnish population: a prospective population based cohort study of the 342 443 FinnGen participants"

**Supplemental file 4.** Results of the time-dependent survival analysis and the sensitivity analysis

### 1. Results of the time-dependent survival analysis

**Supplemental table 1.** Characteristics of FinnGen participants stratified into those who did not sustain an acute event (non-diseased), those who sustained an acute event and either survived the first post acute year (diseased, survived) or died during the first post acute event year (diseased, died during the first year).

| | Non-diseased | Diseased, survived the first year | Diseased, died during the first year | *P*[*] |
|---|---|---|---|---|
| ***Ischemic heart disease*** N | 273 756 | 57 358 | 9 992 | |
| PRS HGS mean (sd) | 0.005 (1.00) | -0.023 (1.00) | -0.006 (1.00) | <0.001 |
| Age at baseline mean (sd) | 64.0 (12.5) | 64.0 (10.7) | 73.5 (10.3) | <0.001[§] |
| BMI mean (sd) | 27.6 (5.41) | 28.0 (4.86) | 27.5 (4.84) | <0.001[§] |
| Current smokers N (%) | 41 168 (25.9) | 13 225 (36.7) | 4 397 (61.4) | <0.001[ǂ] |
| ***Stroke*** N | 231 511 | 34 896 | 5 877 | |
| PRS HGS mean (sd) | 0.008 (1.00) | -0.016 (1.01) | 0.016 (1.00) | <0.001 |
| Age at baseline mean (sd) | 62.8 (12.2) | 65.7 (11.4) | 74.6 (10.6) | <0.001[§] |
| BMI mean (sd) | 27.5 (5.37) | 27.6 (4.84) | 27.1 (4.60) | <0.001[§] |
| Current smokers N (%) | 32 413 (24.1) | 7 935 (35.8) | 1 955 (49.2) | <0.001[ǂ] |
| ***Femur fracture*** N | 326 052 | 6 507 | 2 007 | |
| PRS HGS mean (sd) | -0.001 (1.00) | 0.049 (1.01) | 0.046 (1.00) | <0.001 |
| Age at baseline mean (sd) | 65.9 (12.8) | 71.7 (12.4) | 80.2 (9.19) | <0.001[§] |
| BMI mean (sd) | 27.7 (5.31) | 26.5 (4.85) | 26.6 (4.65) | <0.001[§] |
| Current smokers N (%) | 55 247 (28.7) | 1 821 (42.5) | 777 (53.7) | <0.001[ǂ] |

*One-way analysis of variance, [§]Welch test for variables with unequal variances between groups, [ǂ]Chi square test, PRS HGS = polygenic risk scores for hand grip strength, SD=standard deviation, BMI= body mass index

**Supplemental table 2**. Main effects and interactions of PRS HGS and diseased state on the mortality risk in the FinnGen cohort.

| Phenotype | HR (95% CI) | *P* |
|---|---|---|
| ***Ischemic heart disease*** (N=341 106) | | |
| PRS HGS | **0.97 (0.96–0.99)** | **$2.3 \times 10^{-05}$** |
| Diseased state | | |
| Non-diseased state | 1.00 | |
| First post acute year | **4.71 (4.53–4.90)** | **<0.001** |
| After first post acute year | **1.70 (1.67–1.74)** | **<0.001** |
| Sex | | |
| Women | 1.00 | |
| Men | **1.82 (1.78–1.86)** | **<0.001** |
| PRS HGS and diseased state | | |
| PRS HGS*Non-diseased state | 1.00 | |
| PRS HGS*First post acute year | 1.00 (0.96–1.04) | 0.940 |
| PRS HGS*After first post acute year | 1.00 (0.98–1.02) | 0.960 |
| ***Stroke*** (N=272 284) | | |
| PRS HGS | **0.98 (0.97–0.99)** | **$5.5 \times 10^{-05}$** |
| Diseased state | | |
| Non-diseased state | 1.00 | |
| First post acute year | **5.48 (5.27–5.70)** | **<0.001** |
| After first post acute year | **1.80 (1.76–1.84)** | **<0.001** |
| Sex | | |
| Women | 1.00 | |
| Men | **1.89 (1.85–1.93)** | **<0.001** |
| PRS HGS and diseased state | | |
| PRS HGS*Non-diseased state | 1.00 | |
| PRS HGS*First post acute year | 1.02 (0.98–1.06) | 0.260 |
| PRS HGS*After first post acute year | **0.97 (0.95–0.99)** | **0.004** |
| ***Femur fracture*** (N=334 566) | | |
| PRS HGS | **0.97 (0.96–0.98)** | **$1.8 \times 10^{-10}$** |
| Diseased state | | |
| Non-diseased state | 1.00 | |
| First post acute year | **5.54 (5.24–5.87)** | **<0.001** |
| After first post acute year | **2.22 (2.13–5.87)** | **<0.001** |
| Sex | | |
| Women | | |
| Men | **1.98 (1.93–2.02)** | **<0.001** |
| PRS HGS and diseased state | | |
| PRS HGS*Non-diseased state | 1.00 | |
| PRS HGS*First post acute year | 1.00 (0.95–1.06) | 0.950 |
| PRS HGS*After first post acute year | 0.99 (0.95–1.03) | 0.520 |

Extended Cox regression analysis. Reference group: sex, women; non-diseased state (no acute adverse health event during the follow-up and time before an acute event occurrence of the participants sustaining an acute event). Adjusted for sex, collection year, genotyping batch, and 10 genetic principal components. HR=hazard ratio, CI=confidence interval. Statistically significant values are shown in bold.

### 2. Results of the sensitivity analysis in the FinnGen study

**Supplemental table 3.** Characteristics of participants in the FinnGen study when start of the follow-up was set to the age at the blood sampling for DNA analysis.

| Characteristics | All (N=339 983) | N | Women (N=180 926) | N | Men (N=159 057) | N |
|---|---|---|---|---|---|---|
| Mean (SD) age (y) | 66.30 (12.90) | 339 983 | 64.72 (13.07) | 180 926 | 68.09 (12.47) | 159 057 |
| Mean (SD) BMI (kg/m$^2$) | 27.67 (5.30) | 245 637 | 27.72 (5.85) | 123 128 | 27.62 (4.68) | 122 499 |
| Mean (SD) height (cm) | 170.3 (9.14) | 247 095 | 164.00 (6.31) | 124 043 | 176.70 (6.84) | 123 052 |
| Mean (SD) weight (kg) | 80.48 (17.38) | 251 359 | 74.62 (16.45) | 126 389 | 86.40 (16.26) | 124 970 |
| Smoking status N (%) | | 201 896 | | 99 701 | | 102 195 |
| Never | 96 036 (47.60) | | 60 327 (60.50) | | 35 709 (34.90) | |
| Former | 46 979 (23.30) | | 20 820 (20.88) | | 26 159 (25.60) | |
| Current | 58 881 (29.20) | | 18 554 (18.60) | | 40 327 (39.46) | |

Age at the time of death or at the end of follow-up on 31 December 2021. Phenotype data were obtained from the biobanks (https://www.finngen.fi/en/data_protection/data-protection-statement), SD=standard deviation, BMI=body mass index.

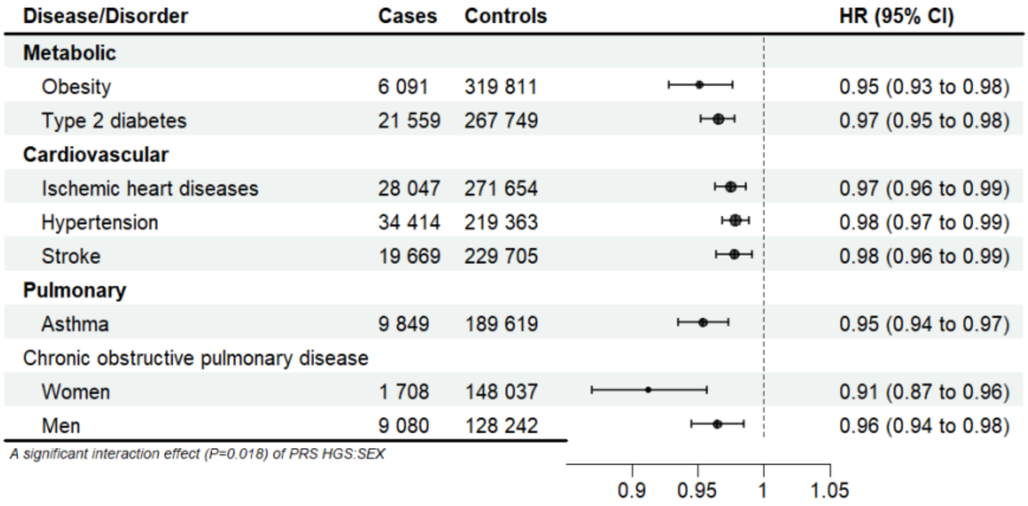

**Supplemental figure 1.** PRS HGS as a predictor of cardiometabolic and pulmonary diseases. Multivariable Cox regression analysis. Adjusted for sex, collection year, genotyping batch, and ten principal genetic components of ancestry. HR=hazard ratio, CI=confidence interval. Note! The start of follow-up from age at the blood sampling for DNA analysis.

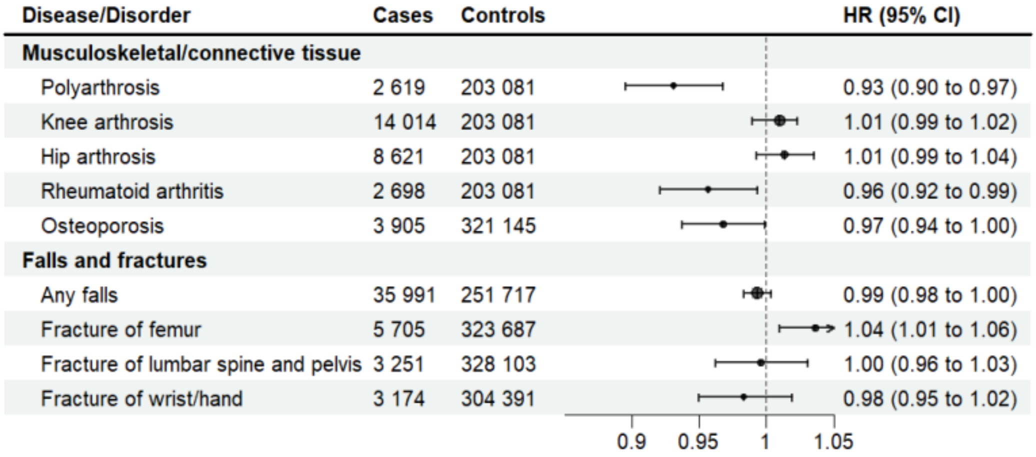

**Supplemental figure 2.** PRS HGS as a predictor of musculoskeletal and connective tissue diseases, falls, and fractures. Multivariable Cox regression analysis. Adjusted for sex, collection year, genotyping batch, and ten principal genetic components of ancestry. HR=hazard ratio, CI=confidence interval. Note! The start of follow-up from age at the blood sampling for DNA analysis.

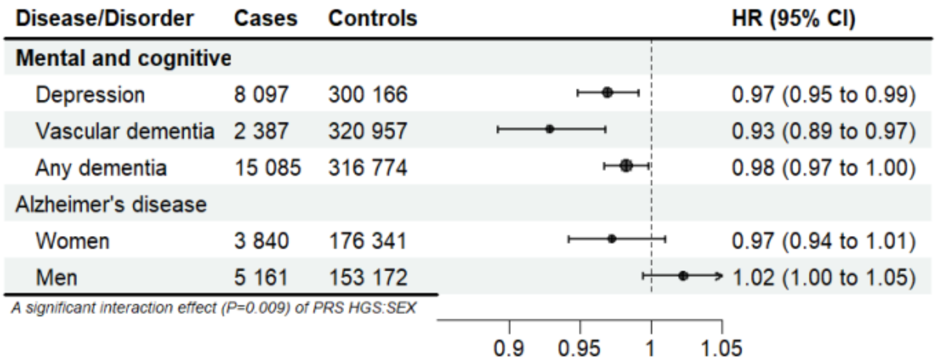

**Supplemental figure 3.** PRS HGS as a predictor of mental and cognitive disorders. Multivariable Cox regression analysis. Adjusted for sex, collection year, genotyping batch, and ten principal genetic components of ancestry. Results with a significant interaction effect (*P* <0.05) between PRS HGS and sex are presented stratified by sex. HR=hazard ratio, CI=confidence interval. Note! The start of follow-up from age at the blood sampling for DNA analysis.

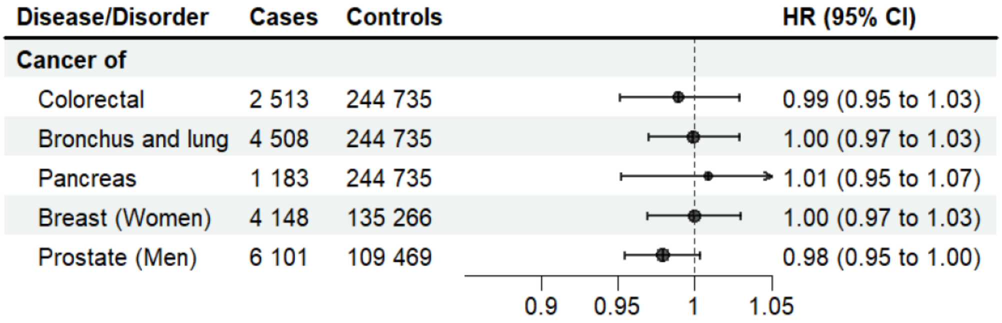

**Supplemental figure 4.** PRS HGS as a predictor of common cancers. Multivariable Cox regression analysis. Adjusted for sex, collection year, genotyping batch, and ten principal genetic components of ancestry. HR=hazard ratio, CI=confidence interval. Note! The start of follow-up from age at the blood sampling for DNA analysis.

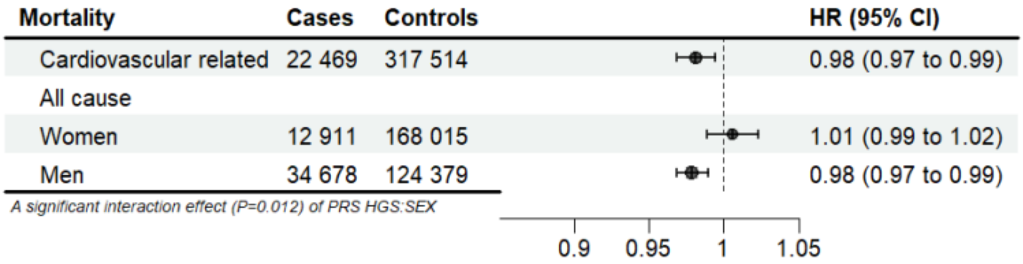

**Supplemental figure 5.** PRS HGS as a predictor of mortality. Multivariable Cox regression analysis. Adjusted for sex, collection year, genotyping batch, and ten principal genetic components of ancestry. Results with a significant interaction effect (*P* <0.05) between PRS HGS and sex are presented stratified by sex. HR=hazard ratio, CI=confidence interval. Note! The start of follow-up from age at the blood sampling for DNA analysis.

### 3. Results of the sensitivity analysis in the FINRISK study

**Supplemental table 4.** Characteristics of participants in the population based FINRISK study.

| Characteristics | All (N=28 543) | N | Women (N=15 138) | N | Men (N=13 405) | N |
|---|---|---|---|---|---|---|
| Mean (SD) age (y) | 67.63 (12.34) | 28 543 | 67.54 (12.57) | 15 138 | 67.73 (12.09) | 13 405 |
| Mean (SD) BMI (kg/m$^2$) | 26.83 (4.70) | 28 448 | 26.50 (5.12) | 15 118 | 27.21 (4.14) | 13 330 |
| Mean (SD) height (cm) | 168.80 (9.32) | 28 452 | 162.60 (6.26) | 15 120 | 175.90 (6.86) | 13 332 |
| Mean (SD) weight (kg) | 76.65 (15.40) | 28 449 | 69.97 (13.52) | 15 119 | 84.22 (13.81) | 13 330 |
| Smoking status N (%) | | 27 449 | | 14 646 | | 12 803 |
| Never | 14 645 (53.35) | | 9 409 (64.24) | | 5 236 (40.90) | |
| Former | 6 267 (22.83) | | 2 461 (16.80) | | 3 806 (29.73) | |
| Current | 6 537 (23.82) | | 2 776 (18.95) | | 3 761 (29.38) | |

Age at the time of death or at the end of follow-up on 31 December 2021. Phenotype data were obtained from the THL Biobank (https://thl.fi/en/web/thlfi-en/research-and-development/research-and-projects/the-national-finrisk-study), SD=standard deviation, BMI=body mass index.

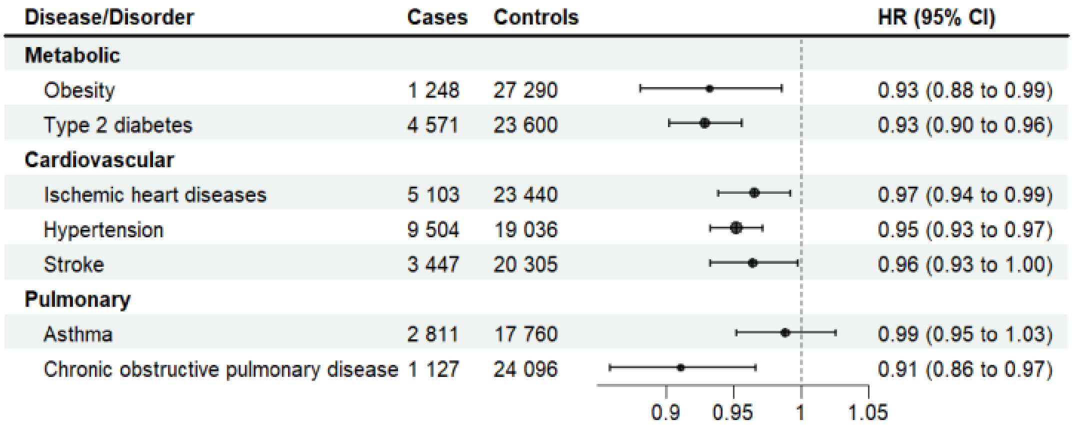

**Supplemental figure 6.** PRS HGS as a predictor of cardiometabolic and pulmonary diseases. Multivariable Cox regression analysis. Adjusted for sex, collection year, genotyping batch, and ten principal genetic components of ancestry. HR=hazard ratio, CI=confidence interval. Note: Start of follow-up from birth, and not from baseline assessment and collection of DNA!

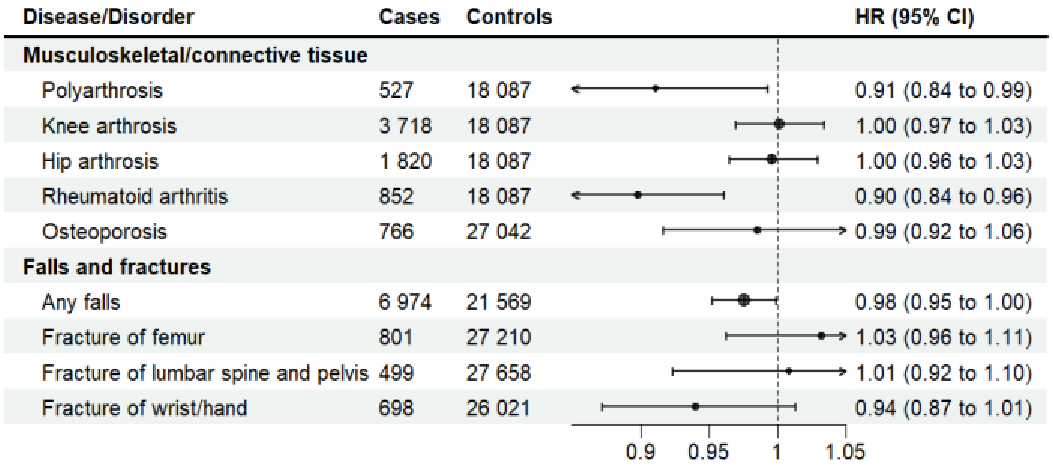

**Supplemental figure 7.** PRS HGS as a predictor of musculoskeletal and connective tissue diseases, falls, and fractures. Multivariable Cox regression analysis. Adjusted for sex, collection year, genotyping batch, and ten principal genetic components of ancestry. HR=hazard ratio, CI=confidence interval. Note: Start of follow-up from birth, and not from baseline assessment and collection of DNA!

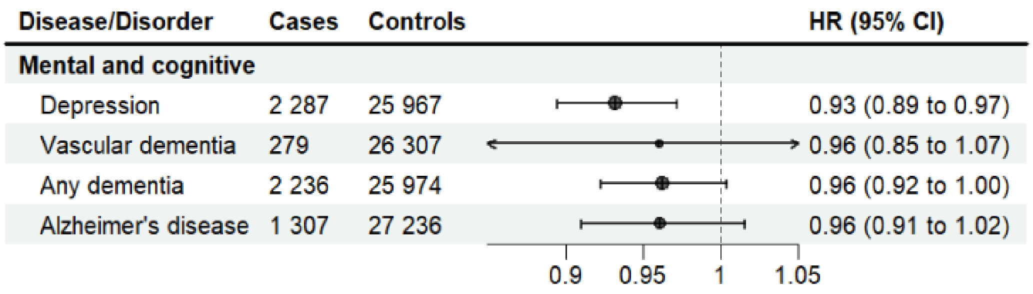

**Supplemental figure 8.** PRS HGS as a predictor of mental and cognitive disorders. Multivariable Cox regression analysis. Adjusted for sex, collection year, genotyping batch, and ten principal genetic components of ancestry. HR=hazard ratio, CI=confidence interval. Note: Start of follow-up from birth, and not from baseline assessment and collection of DNA!

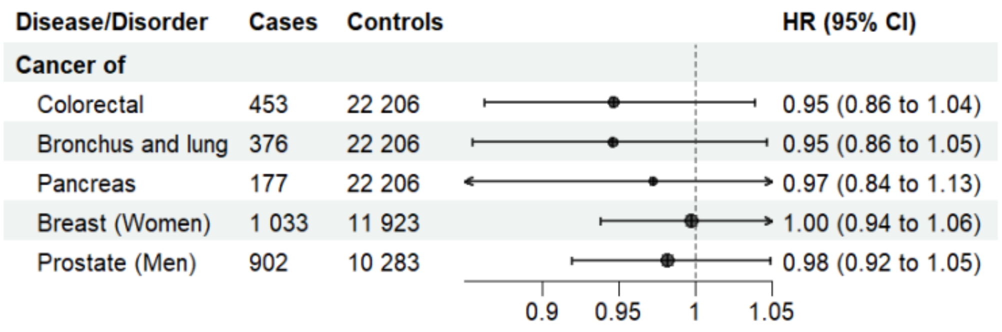

**Supplemental figure 9.** PRS HGS as a predictor of common cancers. Multivariable Cox regression analysis. Adjusted for sex, collection year, genotyping batch, and ten principal genetic components of ancestry. HR=hazard ratio, CI=confidence interval. Note: Start of follow-up from birth, and not from baseline assessment and collection of DNA!

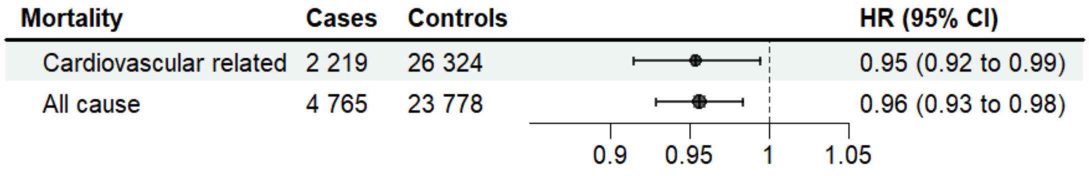

**Supplemental figure 10.** PRS HGS as a predictor of mortality. Multivariable Cox regression analysis. Adjusted for sex, collection year, genotyping batch, and ten principal genetic components of ancestry. HR=hazard ratio, CI=confidence interval. Note: Start of follow-up from birth, and not from baseline assessment and collection of DNA!

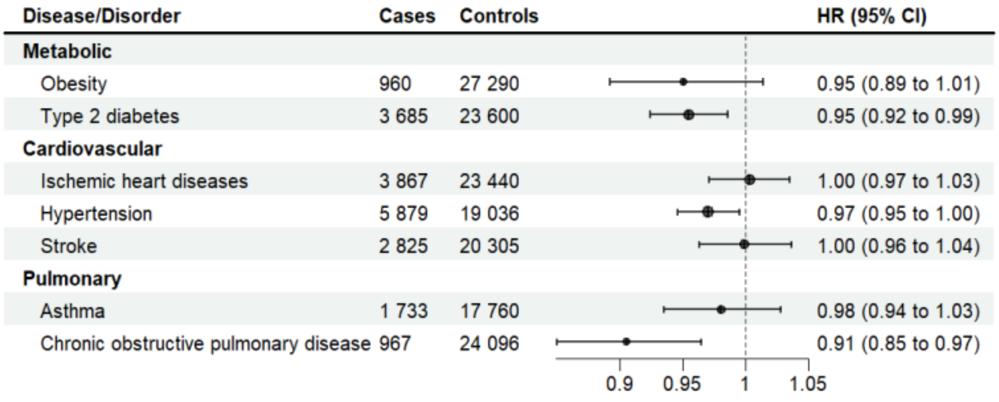

**Supplemental figure 11.** PRS HGS as a predictor of cardiometabolic and pulmonary diseases. Multivariable Cox regression analysis. Adjusted for sex, collection year, genotyping batch, and ten principal genetic components of ancestry. HR=hazard ratio, CI=confidence interval. Note! The start of follow-up from age at baseline data collection, which is also the blood sampling for DNA analysis.

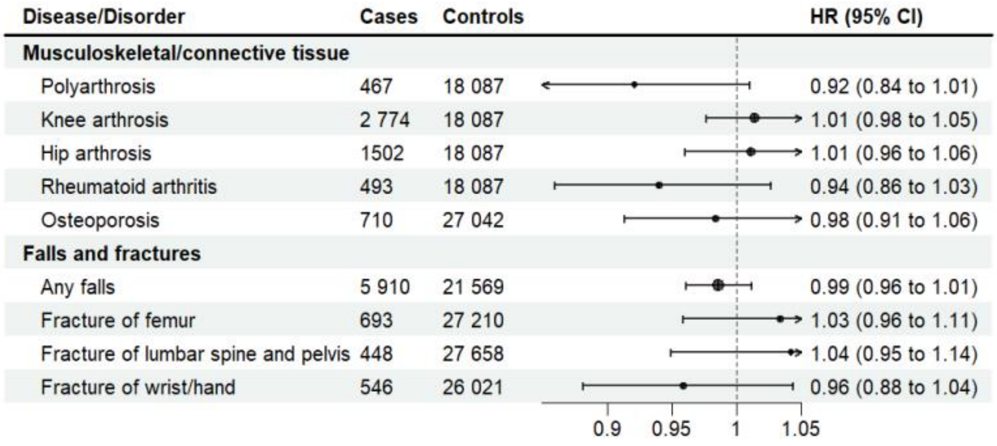

**Supplemental figure 12.** PRS HGS as a predictor of musculoskeletal and connective tissue diseases, falls, and fractures. Multivariable Cox regression analysis. Adjusted for sex, collection year, genotyping batch, and ten principal genetic components of ancestry. HR=hazard ratio, CI=confidence interval. Note! The start of follow-up from age at baseline data collection, which is also the blood sampling for DNA analysis.

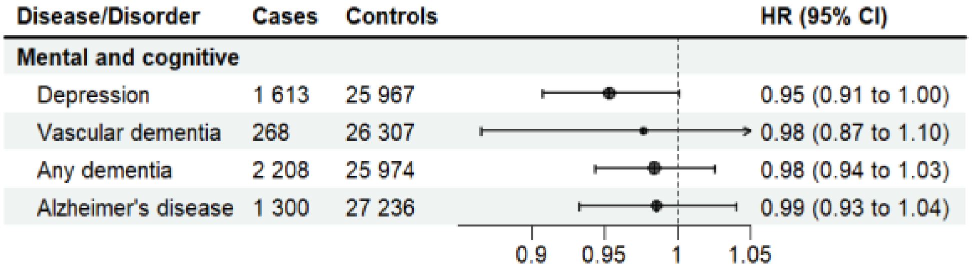

**Supplemental figure 13.** PRS HGS as a predictor of mental and cognitive disorders. Multivariable Cox regression analysis. Adjusted for sex, collection year, genotyping batch, and ten principal genetic components of ancestry. HR=hazard ratio, CI=confidence interval. Note! The start of follow-up from age at baseline data collection, which is also the blood sampling for DNA analysis.

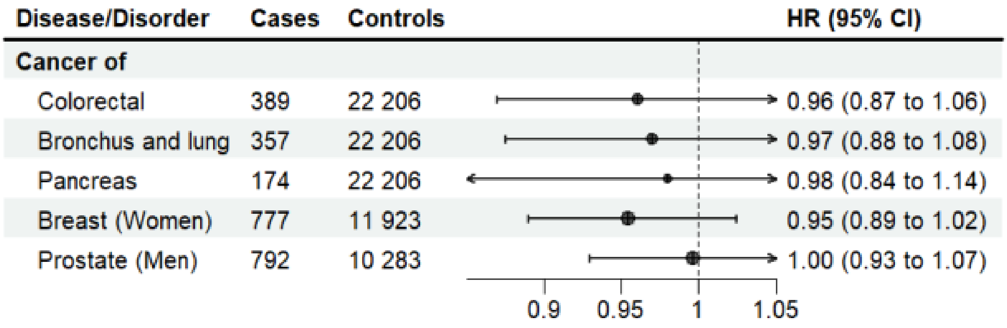

**Supplemental figure 14.** PRS HGS as a predictor of common cancers. Multivariable Cox regression analysis. Adjusted for sex, collection year, genotyping batch, and ten principal genetic components of ancestry. HR=hazard ratio, CI=confidence interval. Note! The start of follow-up from age at baseline data collection, which is also the blood sampling for DNA analysis.

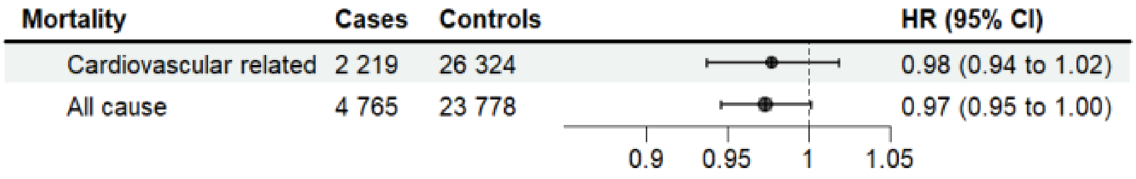

**Supplemental figure 15.** PRS HGS as a predictor of mortality. Multivariable Cox regression analysis. Adjusted for sex, collection year, genotyping batch, and ten principal genetic components of ancestry. HR=hazard ratio, CI=confidence interval. Note! The start of follow-up from age at baseline data collection, which is also the blood sampling for DNA analysis.
