## Supplementary material for "Genome-wide polygenic risk score for muscle strength predicts lower risk for common diseases and longer life span among the Finnish population: a prospective population based cohort study of the 342 443 FinnGen participants": FinnGen-banner_Authors

| Full Name | Affiliation | E-mail | Role 1 | Role 2 |
|---|---|---|---|---|
| Aarno Palotie | Institute for Molecular Medicine Finland (FIMM), HiLIFE, University of Helsinki, Helsinki, Finland; Broad Institute of MIT and Harvard; Massachusetts General Hospital | | Steering Committee | Steering Committee |
| Mark Daly | Institute for Molecular Medicine Finland (FIMM), HiLIFE, University of Helsinki, Helsinki, Finland; Broad Institute of MIT and Harvard; Massachusetts General Hospital | | Steering Committee | Steering Committee |
| Bridget Riley-Gills | Abbvie, Chicago, IL, United States | | Steering Committee | Pharmaceutical companies |
| Howard Jacob | Abbvie, Chicago, IL, United States | | Steering Committee | Pharmaceutical companies |
| Dirk Paul | Astra Zeneca, Cambridge, United Kingdom | | Steering Committee | Pharmaceutical companies |
| Slavé Petrovski | Astra Zeneca, Cambridge, United Kingdom | | Steering Committee | Pharmaceutical companies |
| Heiko Runz | Biogen, Cambridge, MA, United States | | Steering Committee | Pharmaceutical companies |
| Sally John | Biogen, Cambridge, MA, United States | | Steering Committee | Pharmaceutical companies |
| George Okafo | Boehringer Ingelheim, Ingelheim am Rhein, Germany | | Steering Committee | Pharmaceutical companies |
| Nathan Lawless | Boehringer Ingelheim, Ingelheim am Rhein, Germany | | Steering Committee | Pharmaceutical companies |
| Heli Salminen-Mankonen | Boehringer Ingelheim, Ingelheim am Rhein, Germany | heli.salminen-mankonen@boehringer-ingelheir | Steering Committee | Pharmaceutical companies |
| Robert Plenge | Bristol Myers Squibb, New York, NY, United States | | Steering Committee | Pharmaceutical companies |
| Joseph Maranville | Bristol Myers Squibb, New York, NY, United States | | Steering Committee | Pharmaceutical companies |
| Mark McCarthy | Genentech, San Francisco, CA, United States | | Steering Committee | Pharmaceutical companies |
| Margaret G. Ehm | GlaxoSmithKline, Collegeville, PA, United States | | Steering Committee | Pharmaceutical companies |
| Kirsi Auro | GlaxoSmithKline, Espoo, Finland | | Steering Committee | Pharmaceutical companies |
| Simonne Longerich | Merck, Kenilworth, NJ, United States | | Steering Committee | Pharmaceutical companies |
| Anders Mälarstig | Pfizer, New York, NY, United States | | Steering Committee | Pharmaceutical companies |
| Katherine Klinger | Translational Sciences, Sanofi R&D, Framingham, MA, USA | | Steering Committee | Pharmaceutical companies |
| Clement Chatelain | Translational Sciences, Sanofi R&D, Framingham, MA, USA | | Steering Committee | Pharmaceutical companies |
| Matthias Gossel | Translational Sciences, Sanofi R&D, Framingham, MA, USA | | Steering Committee | Pharmaceutical companies |
| Karol Estrada | Maze Therapeutics, San Francisco, CA, United States | | Steering Committee | Pharmaceutical companies |
| Robert Graham | Maze Therapeutics, San Francisco, CA, United States | | Steering Committee | Pharmaceutical companies |
| Robert Yang | Janssen Biotech, Beerse, Belgium | | Steering Committee | Pharmaceutical companies |
| Chris O´Donnell | Novartis Institutes for BioMedical Research, Cambridge, MA, United States | | Steering Committee | Pharmaceutical companies |
| Tomi P. Mäkelä | HiLIFE, University of Helsinki, Finland, Finland | | Steering Committee | University of Helsinki & Biobanks |
| Jaakko Kaprio | Institute for Molecular Medicine Finland (FIMM), HiLIFE, University of Helsinki, Helsinki, Finland | | Steering Committee | University of Helsinki & Biobanks |
| Petri Virolainen | Auria Biobank / University of Turku / Hospital District of Southwest Finland, Turku, Finland | | Steering Committee | University of Helsinki & Biobanks |
| Antti Hakanen | Auria Biobank / University of Turku / Hospital District of Southwest Finland, Turku, Finland | | Steering Committee | University of Helsinki & Biobanks |
| Terhi Kilpi | THL Biobank / Finnish Institute for Health and Welfare (THL), Helsinki, Finland | | Steering Committee | University of Helsinki & Biobanks |
| Markus Perola | THL Biobank / Finnish Institute for Health and Welfare (THL), Helsinki, Finland | | Steering Committee | University of Helsinki & Biobanks |
| Jukka Partanen | Finnish Red Cross Blood Service / Finnish Hematology Registry and Clinical Biobank, Helsinki, Finland | | Steering Committee | University of Helsinki & Biobanks |
| Anne Pitkäranta | Helsinki Biobank / Helsinki University and Hospital District of Helsinki and Uusimaa, Helsinki | | Steering Committee | University of Helsinki & Biobanks |
| Taneli Raivio | Helsinki Biobank / Helsinki University and Hospital District of Helsinki and Uusimaa, Helsinki | | Steering Committee | University of Helsinki & Biobanks |
| Jani Tikkanen | Northern Finland Biobank Borealis / University of Oulu / Northern Ostrobothnia Hospital District, Oulu, Finland | | Steering Committee | University of Helsinki & Biobanks |
| Raisa Serpi | Northern Finland Biobank Borealis / University of Oulu / Northern Ostrobothnia Hospital District, Oulu, Finland | | Steering Committee | University of Helsinki & Biobanks |
| Tarja Laitinen | Finnish Clinical Biobank Tampere / University of Tampere / Pirkanmaa Hospital District, Tampere, Finland | | Steering Committee | University of Helsinki & Biobanks |
| Veli-Matti Kosma | Biobank of Eastern Finland / University of Eastern Finland / Northern Savo Hospital District, Kuopio, Finland | | Steering Committee | University of Helsinki & Biobanks |
| Jari Laukkanen | Central Finland Biobank / University of Jyväskylä / Central Finland Health Care District, Jyväskylä, Finland | | Steering Committee | University of Helsinki & Biobanks |
| Marco Hautalahti | FINBB - Finnish biobank cooperative | | Steering Committee | University of Helsinki & Biobanks |
| Outi Tuovila | Business Finland, Helsinki, Finland | | Steering Committee | Other Experts/ Non-Voting Members |
| Raimo Pakkanen | Business Finland, Helsinki, Finland | | Steering Committee | Other Experts/ Non-Voting Members |
| Jeffrey Waring | Abbvie, Chicago, IL, United States | | Scientific Committee | Pharmaceutical companies |
| Bridget Riley-Gillis | Abbvie, Chicago, IL, United States | | Scientific Committee | Pharmaceutical companies |
| Fedik Rahimov | Abbvie, Chicago, IL, United States | | Scientific Committee | Pharmaceutical companies |
| Ioanna Tachmazidou | Astra Zeneca, Cambridge, United Kingdom | | Scientific Committee | Pharmaceutical companies |
| Chia-Yen Chen | Biogen, Cambridge, MA, United States | | Scientific Committee | Pharmaceutical companies |
| Heiko Runz | Biogen, Cambridge, MA, United States | | Scientific Committee | Pharmaceutical companies |
| Zhihao Ding | Boehringer Ingelheim, Ingelheim am Rhein, Germany | | Scientific Committee | Pharmaceutical companies |
| Marc Jung | Boehringer Ingelheim, Ingelheim am Rhein, Germany | | Scientific Committee | Pharmaceutical companies |
| Shameek Biswas | Bristol Myers Squibb, New York, NY, United States | | Scientific Committee | Pharmaceutical companies |
| Rion Pendergrass | Genentech, San Francisco, CA, United States | | Scientific Committee | Pharmaceutical companies |
| Margaret G. Ehm | GlaxoSmithKline, Collegeville, PA, United States | | Scientific Committee | Pharmaceutical companies |
| David Pulford | GlaxoSmithKline, Stevenage, United Kingdom | | Scientific Committee | Pharmaceutical companies |
| Neha Raghavan | Merck, Kenilworth, NJ, United States | | Scientific Committee | Pharmaceutical companies |
| Adriana Huertas-Vazquez | Merck, Kenilworth, NJ, United States | | Scientific Committee | Pharmaceutical companies |
| Jae-Hoon Sul | Merck, Kenilworth, NJ, United States | | Scientific Committee | Pharmaceutical companies |
| Anders Mälarstig | Pfizer, New York, NY, United States | | Scientific Committee | Pharmaceutical companies |
| Xinli Hu | Pfizer, New York, NY, United States | | Scientific Committee | Pharmaceutical companies |
| Åsa Hedman | Pfizer, New York, NY, United States | | Scientific Committee | Pharmaceutical companies |
| Katherine Klinger | Translational Sciences, Sanofi R&D, Framingham, MA, USA | | Scientific Committee | Pharmaceutical companies |
| Robert Graham | Maze Therapeutics, San Francisco, CA, United States | | Scientific Committee | Pharmaceutical companies |
| Manuel Rivas | Maze Therapeutics, San Francisco, CA, United States | | Scientific Committee | Pharmaceutical companies |
| Dawn Waterworth | Janssen Research & Development, LLC, Spring House, PA, United States | | Scientific Committee | Pharmaceutical companies |
| Nicole Renaud | Novartis Institutes for BioMedical Research, Cambridge, MA, United States | | Scientific Committee | Pharmaceutical companies |
| Ma´en Obeidat | Novartis Institutes for BioMedical Research, Cambridge, MA, United States | | Scientific Committee | Pharmaceutical companies |
| Samuli Ripatti | Institute for Molecular Medicine Finland (FIMM), HiLIFE, University of Helsinki, Helsinki, Finland | | Scientific Committee | University of Helsinki & Biobanks |
| Johanna Schleutker | Auria Biobank / Univ. of Turku / Hospital District of Southwest Finland, Turku, Finland | | Scientific Committee | University of Helsinki & Biobanks |
| Markus Perola | THL Biobank / Finnish Institute for Health and Welfare (THL), Helsinki, Finland | | Scientific Committee | University of Helsinki & Biobanks |
| Mikko Arvas | Finnish Red Cross Blood Service / Finnish Hematology Registry and Clinical Biobank, Helsinki, Finland | | Scientific Committee | University of Helsinki & Biobanks |
| Olli Carpén | Helsinki Biobank / Helsinki University and Hospital District of Helsinki and Uusimaa, Helsinki | | Scientific Committee | University of Helsinki & Biobanks |
| Reetta Hinttala | Northern Finland Biobank Borealis / University of Oulu / Northern Ostrobothnia Hospital District, Oulu, Finland | | Scientific Committee | University of Helsinki & Biobanks |
| Johannes Kettunen | Northern Finland Biobank Borealis / University of Oulu / Northern Ostrobothnia Hospital District, Oulu, Finland | | Scientific Committee | University of Helsinki & Biobanks |
| Arto Mannermaa | Biobank of Eastern Finland / University of Eastern Finland / Northern Savo Hospital District, Kuopio, Finland | | Scientific Committee | University of Helsinki & Biobanks |
| Katriina Aalto-Setälä | Faculty of Medicine and Health Technology, Tampere University, Tampere, Finland | | Scientific Committee | University of Helsinki & Biobanks |
| Mika Kähönen | Finnish Clinical Biobank Tampere / University of Tampere / Pirkanmaa Hospital District, Tampere, Finland | | Scientific Committee | University of Helsinki & Biobanks |
| Jari Laukkanen | Central Finland Biobank / University of Jyväskylä / Central Finland Health Care District, Jyväskylä, Finland | | Scientific Committee | University of Helsinki & Biobanks |
| Johanna Mäkelä | FINBB - Finnish biobank cooperative | | Scientific Committee | University of Helsinki & Biobanks |
| Reetta Kälviäinen | Northern Savo Hospital District, Kuopio, Finland | | Clinical Groups | Neurology Group |
| Valtteri Julkunen | Northern Savo Hospital District, Kuopio, Finland | | Clinical Groups | Neurology Group |
| Hilkka Soininen | Northern Savo Hospital District, Kuopio, Finland | | Clinical Groups | Neurology Group |
| Anne Remes | Northern Ostrobothnia Hospital District, Oulu, Finland | | Clinical Groups | Neurology Group |
| Mikko Hiltunen | University of Eastern Finland, Kuopio, Finland | | Clinical Groups | Neurology Group |
| Jukka Peltola | Pirkanmaa Hospital District, Tampere, Finland | | Clinical Groups | Neurology Group |
| Minna Raivio | Hospital District of Helsinki and Uusimaa, Helsinki, Finland | | Clinical Groups | Neurology Group |
| Pentti Tienari | Hospital District of Helsinki and Uusimaa, Helsinki, Finland | | Clinical Groups | Neurology Group |
| Juha Rinne | Hospital District of Southwest Finland, Turku, Finland | | Clinical Groups | Neurology Group |
| Roosa Kallionpää | Hospital District of Southwest Finland, Turku, Finland | | Clinical Groups | Neurology Group |
| Juulia Partanen | Institute for Molecular Medicine Finland, HiLIFE, University of Helsinki, Finland | | Clinical Groups | Neurology Group |
| Ali Abbasi | Abbvie, Chicago, IL, United States | | Clinical Groups | Neurology Group |
| Adam Ziemann | Abbvie, Chicago, IL, United States | | Clinical Groups | Neurology Group |
| Nizar Smaoui | Abbvie, Chicago, IL, United States | | Clinical Groups | Neurology Group |
| Anne Lehtonen | Abbvie, Chicago, IL, United States | | Clinical Groups | Neurology Group |

| | | | | |
|---|---|---|---|---|
| Susan Eaton | Biogen, Cambridge, MA, United States | | Clinical Groups | **Neurology Group** |
| Heiko Runz | Biogen, Cambridge, MA, United States | | Clinical Groups | **Neurology Group** |
| Sanni Lahdenperä | Biogen, Cambridge, MA, United States | | Clinical Groups | **Neurology Group** |
| Shameek Biswas | Bristol Myers Squibb, New York, NY, United States | | Clinical Groups | **Neurology Group** |
| Natalie Bowers | Genentech, San Francisco, CA, United States | | Clinical Groups | **Neurology Group** |
| Edmond Teng | Genentech, San Francisco, CA, United States | | Clinical Groups | **Neurology Group** |
| Rion Pendergrass | Genentech, San Francisco, CA, United States | | Clinical Groups | **Neurology Group** |
| Fanli Xu | GlaxoSmithKline, Brentford, United Kingdom | | Clinical Groups | **Neurology Group** |
| David Pulford | GlaxoSmithKline, Stevenage, United Kingdom | | Clinical Groups | **Neurology Group** |
| Kirsi Auro | GlaxoSmithKline, Espoo, Finland | | Clinical Groups | **Neurology Group** |
| Laura Addis | GlaxoSmithKline, Brentford, United Kingdom | | Clinical Groups | **Neurology Group** |
| John Eicher | GlaxoSmithKline, Brentford, United Kingdom | | Clinical Groups | **Neurology Group** |
| Qingqin S Li | Janssen Research & Development, LLC, Titusville, NJ 08560, United States | | Clinical Groups | **Neurology Group** |
| Karen He | Janssen Research & Development, LLC, Spring House, PA, United States | | Clinical Groups | **Neurology Group** |
| Ekaterina Khramtsova | Janssen Research & Development, LLC, Spring House, PA, United States | | Clinical Groups | **Neurology Group** |
| Neha Raghavan | Merck, Kenilworth, NJ, United States | | Clinical Groups | **Neurology Group** |
| Martti Färkkilä | Hospital District of Helsinki and Uusimaa, Helsinki, Finland | | Clinical Groups | **Gastroenterology Group** |
| Jukka Koskela | Hospital District of Helsinki and Uusimaa, Helsinki, Finland | | Clinical Groups | **Gastroenterology Group** |
| Sampsa Pikkarainen | Hospital District of Helsinki and Uusimaa, Helsinki, Finland | | Clinical Groups | **Gastroenterology Group** |
| Airi Jussila | Pirkanmaa Hospital District, Tampere, Finland | | Clinical Groups | **Gastroenterology Group** |
| Katri Kaukinen | Pirkanmaa Hospital District, Tampere, Finland | | Clinical Groups | **Gastroenterology Group** |
| Timo Blomster | Northern Ostrobothnia Hospital District, Oulu, Finland | | Clinical Groups | **Gastroenterology Group** |
| Mikko Kiviniemi | Northern Savo Hospital District, Kuopio, Finland | | Clinical Groups | **Gastroenterology Group** |
| Markku Voutilainen | Hospital District of Southwest Finland, Turku, Finland | | Clinical Groups | **Gastroenterology Group** |
| Mark Daly | Institute for Molecular Medicine, Finland (FIMM), HiLIFE, University of Helsinki, Helsinki, Finland; Broad Institute of MIT and Harvard; Massachusetts General Hospital | | Clinical Groups | **Gastroenterology Group** |
| Ali Abbasi | Abbvie, Chicago, IL, United States | | Clinical Groups | **Gastroenterology Group** |
| Jeffrey Waring | Abbvie, Chicago, IL, United States | | Clinical Groups | **Gastroenterology Group** |
| Nizar Smaoui | Abbvie, Chicago, IL, United States | | Clinical Groups | **Gastroenterology Group** |
| Fedik Rahimov | Abbvie, Chicago, IL, United States | | Clinical Groups | **Gastroenterology Group** |
| Anne Lehtonen | Abbvie, Chicago, IL, United States | | Clinical Groups | **Gastroenterology Group** |
| Tim Lu | Genentech, San Francisco, CA, United States | | Clinical Groups | **Gastroenterology Group** |
| Natalie Bowers | Genentech, San Francisco, CA, United States | | Clinical Groups | **Gastroenterology Group** |
| Rion Pendergrass | Genentech, San Francisco, CA, United States | | Clinical Groups | **Gastroenterology Group** |
| Linda McCarthy | GlaxoSmithKline, Brentford, United Kingdom | | Clinical Groups | **Gastroenterology Group** |
| Amy Hart | Janssen Research & Development, LLC, Spring House, PA, United States | | Clinical Groups | **Gastroenterology Group** |
| Meijian Guan | Janssen Research & Development, LLC, Spring House, PA, United States | | Clinical Groups | **Gastroenterology Group** |
| Jason Miller | Merck, Kenilworth, NJ, United States | | Clinical Groups | **Gastroenterology Group** |
| Kirsi Kalpala | Pfizer, New York, NY, United States | | Clinical Groups | **Gastroenterology Group** |
| Melissa Miller | Pfizer, New York, NY, United States | | Clinical Groups | **Gastroenterology Group** |
| Xinli Hu | Pfizer, New York, NY, United States | | Clinical Groups | **Gastroenterology Group** |
| Kari Eklund | Hospital District of Helsinki and Uusimaa, Helsinki, Finland | | Clinical Groups | **Rheumatology Group** |
| Antti Palomäki | Hospital District of Southwest Finland, Turku, Finland | | Clinical Groups | **Rheumatology Group** |
| Pia Isomäki | Pirkanmaa Hospital District, Tampere, Finland | | Clinical Groups | **Rheumatology Group** |
| Laura Pirilä | Hospital District of Southwest Finland, Turku, Finland | | Clinical Groups | **Rheumatology Group** |
| Oili Kaipiainen-Seppänen | Northern Savo Hospital District, Kuopio, Finland | | Clinical Groups | **Rheumatology Group** |
| Johanna Huhtakangas | Northern Ostrobothnia Hospital District, Oulu, Finland | | Clinical Groups | **Rheumatology Group** |
| Nina Mars | Institute for Molecular Medicine Finland (FIMM), HiLIFE, University of Helsinki, Helsinki, Finland | | Clinical Groups | **Rheumatology Group** |
| Ali Abbasi | Abbvie, Chicago, IL, United States | | Clinical Groups | **Rheumatology Group** |
| Jeffrey Waring | Abbvie, Chicago, IL, United States | | Clinical Groups | **Rheumatology Group** |
| Fedik Rahimov | Abbvie, Chicago, IL, United States | | Clinical Groups | **Rheumatology Group** |
| Apinya Lertratanakul | Abbvie, Chicago, IL, United States | | Clinical Groups | **Rheumatology Group** |
| Nizar Smaoui | Abbvie, Chicago, IL, United States | | Clinical Groups | **Rheumatology Group** |
| Anne Lehtonen | Abbvie, Chicago, IL, United States | | Clinical Groups | **Rheumatology Group** |
| Coralie Viollet | AstraZeneca, Cambridge, United Kingdom | | Clinical Groups | **Rheumatology Group** |
| Marla Hochfeld | Bristol Myers Squibb, New York, NY, United States | | Clinical Groups | **Rheumatology Group** |
| Natalie Bowers | Genentech, San Francisco, CA, United States | | Clinical Groups | **Rheumatology Group** |
| Rion Pendergrass | Genentech, San Francisco, CA, United States | | Clinical Groups | **Rheumatology Group** |
| Jorge Esparza Gordillo | GlaxoSmithKline, Brentford, United Kingdom | | Clinical Groups | **Rheumatology Group** |
| Kirsi Auro | GlaxoSmithKline, Espoo, Finland | | Clinical Groups | **Rheumatology Group** |
| Dawn Waterworth | Janssen Research & Development, LLC, Spring House, PA, United States | | Clinical Groups | **Rheumatology Group** |
| Fabiana Farias | Merck, Kenilworth, NJ, United States | | Clinical Groups | **Rheumatology Group** |
| Kirsi Kalpala | Pfizer, New York, NY, United States | | Clinical Groups | **Rheumatology Group** |
| Nan Bing | Pfizer, New York, NY, United States | | Clinical Groups | **Rheumatology Group** |
| Xinli Hu | Pfizer, New York, NY, United States | | Clinical Groups | **Rheumatology Group** |
| Tarja Laitinen | Pirkanmaa Hospital District, Tampere, Finland | | Clinical Groups | **Pulmonology Group** |
| Margit Pelkonen | Northern Savo Hospital District, Kuopio, Finland | | Clinical Groups | **Pulmonology Group** |
| Paula Kauppi | Hospital District of Helsinki and Uusimaa, Helsinki, Finland | | Clinical Groups | **Pulmonology Group** |
| Hannu Kankaanranta | University of Gothenburg, Gothenburg, Sweden/ Seinäjoki Central Hospital, Seinäjoki, Finland/ Tampere University, Tampere, Finland | | Clinical Groups | **Pulmonology Group** |
| Terttu Harju | Northern Ostrobothnia Hospital District, Oulu, Finland | | Clinical Groups | **Pulmonology Group** |
| Riitta Lahesmaa | Hospital District of Southwest Finland, Turku, Finland | | Clinical Groups | **Pulmonology Group** |
| Nizar Smaoui | Abbvie, Chicago, IL, United States | | Clinical Groups | **Pulmonology Group** |
| Coralie Viollet | AstraZeneca, Cambridge, United Kingdom | | Clinical Groups | **Pulmonology Group** |
| Susan Eaton | Biogen, Cambridge, MA, United States | | Clinical Groups | **Pulmonology Group** |
| Hubert Chen | Genentech, San Francisco, CA, United States | | Clinical Groups | **Pulmonology Group** |
| Rion Pendergrass | Genentech, San Francisco, CA, United States | | Clinical Groups | **Pulmonology Group** |
| Natalie Bowers | Genentech, San Francisco, CA, United States | | Clinical Groups | **Pulmonology Group** |
| Joanna Betts | GlaxoSmithKline, Brentford, United Kingdom | | Clinical Groups | **Pulmonology Group** |
| Kirsi Auro | GlaxoSmithKline, Espoo, Finland | | Clinical Groups | **Pulmonology Group** |
| Rajashree Mishra | GlaxoSmithKline, Brentford, United Kingdom | | Clinical Groups | **Pulmonology Group** |
| Majd Mouded | Novartis, Basel, Switzerland | | Clinical Groups | **Pulmonology Group** |
| Debby Ngo | Novartis, Basel, Switzerland | | Clinical Groups | **Pulmonology Group** |
| Teemu Niiranen | Finnish Institute for Health and Welfare (THL), Helsinki, Finland | | Clinical Groups | **Cardiometabolic Diseases Group** |
| Felix Vaura | Finnish Institute for Health and Welfare (THL), Helsinki, Finland | | Clinical Groups | **Cardiometabolic Diseases Group** |
| Veikko Salomaa | Finnish Institute for Health and Welfare (THL), Helsinki, Finland | | Clinical Groups | **Cardiometabolic Diseases Group** |
| Kaj Metsärinne | Hospital District of Southwest Finland, Turku, Finland | | Clinical Groups | **Cardiometabolic Diseases Group** |
| Jenni Aittokallio | Hospital District of Southwest Finland, Turku, Finland | | Clinical Groups | **Cardiometabolic Diseases Group** |
| Mika Kähönen | Pirkanmaa Hospital District, Tampere, Finland | | Clinical Groups | **Cardiometabolic Diseases Group** |
| Jussi Hernesniemi | Pirkanmaa Hospital District, Tampere, Finland | | Clinical Groups | **Cardiometabolic Diseases Group** |
| Daniel Gordin | Hospital District of Helsinki and Uusimaa, Helsinki, Finland | | Clinical Groups | **Cardiometabolic Diseases Group** |
| Juha Sinisalo | Hospital District of Helsinki and Uusimaa, Helsinki, Finland | | Clinical Groups | **Cardiometabolic Diseases Group** |
| Marja-Riitta Taskinen | Hospital District of Helsinki and Uusimaa, Helsinki, Finland | | Clinical Groups | **Cardiometabolic Diseases Group** |
| Tiinamaija Tuomi | Hospital District of Helsinki and Uusimaa, Helsinki, Finland | | Clinical Groups | **Cardiometabolic Diseases Group** |
| Timo Hiltunen | Hospital District of Helsinki and Uusimaa, Helsinki, Finland | | Clinical Groups | **Cardiometabolic Diseases Group** |
| Jari Laukkanen | Central Finland Health Care District, Jyväskylä, Finland | | Clinical Groups | **Cardiometabolic Diseases Group** |
| Amanda Elliott | Institute for Molecular Medicine Finland (FIMM), HiLIFE, University of Helsinki, Helsinki, Finland; Broad Institute, Cambridge, MA, USA and Massachusetts General Hospital, Boston, MA, USA | | Clinical Groups | **Cardiometabolic Diseases Group** |
| Mary Pat Reeve | Institute for Molecular Medicine Finland (FIMM), HiLIFE, University of Helsinki, Helsinki, Finland | | Clinical Groups | **Cardiometabolic Diseases Group** |
| Sanni Ruotsalainen | Institute for Molecular Medicine Finland (FIMM), HiLIFE, University of Helsinki, Helsinki, Finland | | Clinical Groups | **Cardiometabolic Diseases Group** |
| Dirk Paul | Astra Zeneca, Cambridge, United Kingdom | | Clinical Groups | **Cardiometabolic Diseases Group** |
| Natalie Bowers | Genentech, San Francisco, CA, United States | | Clinical Groups | **Cardiometabolic Diseases Group** |
| Rion Pendergrass | Genentech, San Francisco, CA, United States | | Clinical Groups | **Cardiometabolic Diseases Group** |
| Audrey Chu | GlaxoSmithKline, Brentford, United Kingdom | | Clinical Groups | **Cardiometabolic Diseases Group** |
| Kirsi Auro | GlaxoSmithKline, Espoo, Finland | | Clinical Groups | **Cardiometabolic Diseases Group** |

| | | | | |
|---|---|---|---|---|
| Dermot Reilly | Janssen Research & Development, LLC, Boston, MA, United States | | Clinical Groups | **Cardiometabolic Diseases Group** |
| Mike Mendelson | Novartis, Boston, MA, United States | | Clinical Groups | **Cardiometabolic Diseases Group** |
| Jaakko Parkkinen | Pfizer, New York, NY, United States | | Clinical Groups | **Cardiometabolic Diseases Group** |
| Melissa Miller | Pfizer, New York, NY, United States | | Clinical Groups | **Cardiometabolic Diseases Group** |
| Tuomo Meretoja | Hospital District of Helsinki and Uusimaa, Helsinki, Finland | | Clinical Groups | **Oncology Group** |
| Heikki Joensuu | Hospital District of Helsinki and Uusimaa, Helsinki, Finland | | Clinical Groups | **Oncology Group** |
| Olli Carpén | Hospital District of Helsinki and Uusimaa, Helsinki, Finland | | Clinical Groups | **Oncology Group** |
| Johanna Mattson | Hospital District of Helsinki and Uusimaa, Helsinki, Finland | | Clinical Groups | **Oncology Group** |
| Eveliina Salminen | Hospital District of Helsinki and Uusimaa, Helsinki, Finland | | Clinical Groups | **Oncology Group** |
| Annika Auranen | Pirkanmaa Hospital District , Tampere, Finland | | Clinical Groups | **Oncology Group** |
| Peeter Karihtala | Northern Ostrobothnia Hospital District, Oulu, Finland | | Clinical Groups | **Oncology Group** |
| Päivi Auvinen | Northern Savo Hospital District, Kuopio, Finland | | Clinical Groups | **Oncology Group** |
| Klaus Elenius | Hospital District of Southwest Finland, Turku, Finland | | Clinical Groups | **Oncology Group** |
| Johanna Schleutker | Hospital District of Southwest Finland, Turku, Finland | | Clinical Groups | **Oncology Group** |
| Esa Pitkänen | Institute for Molecular Medicine Finland (FIMM), HiLIFE, University of Helsinki, Helsinki, Finland | | Clinical Groups | **Oncology Group** |
| Nina Mars | Institute for Molecular Medicine Finland (FIMM), HiLIFE, University of Helsinki, Helsinki, Finland | | Clinical Groups | **Oncology Group** |
| Mark Daly | Institute for Molecular Medicine Finland (FIMM), HiLIFE, University of Helsinki, Helsinki, Finland; Broad Institute of MIT and Harvard; Massachusetts General Hospital | | Clinical Groups | **Oncology Group** |
| Relja Popovic | Abbvie, Chicago, IL, United States | | Clinical Groups | **Oncology Group** |
| Jeffrey Waring | Abbvie, Chicago, IL, United States | | Clinical Groups | **Oncology Group** |
| Bridget Riley-Gillis | Abbvie, Chicago, IL, United States | | Clinical Groups | **Oncology Group** |
| Anne Lehtonen | Abbvie, Chicago, IL, United States | | Clinical Groups | **Oncology Group** |
| Margarete Fabre | AstraZeneca, Cambridge, United Kingdom | | Clinical Groups | **Oncology Group** |
| Jennifer Schutzman | Genentech, San Francisco, CA, United States | | Clinical Groups | **Oncology Group** |
| Natalie Bowers | Genentech, San Francisco, CA, United States | | Clinical Groups | **Oncology Group** |
| Rion Pendergrass | Genentech, San Francisco, CA, United States | | Clinical Groups | **Oncology Group** |
| Diptee Kulkarni | GlaxoSmithKline, Brentford, United Kingdom | | Clinical Groups | **Oncology Group** |
| Kirsi Auro | GlaxoSmithKline, Espoo, Finland | | Clinical Groups | **Oncology Group** |
| Alessandro Porello | Janssen Research & Development, LLC, Spring House, PA, United States | | Clinical Groups | **Oncology Group** |
| Andrey Loboda | Merck, Kenilworth, NJ, United States | | Clinical Groups | **Oncology Group** |
| Heli Lehtonen | Pfizer, New York, NY, United States | | Clinical Groups | **Oncology Group** |
| Stefan McDonough | Pfizer, New York, NY, United States | | Clinical Groups | **Oncology Group** |
| Sauli Vuoti | Janssen-Cilag Oy, Espoo, Finland | | Clinical Groups | **Oncology Group** |
| Kai Kaarniranta | Northern Savo Hospital District, Kuopio, Finland; Department of Molecular Genetics, University of Lodz, Lodz, Poland | | Clinical Groups | **Opthalmology Group** |
| Joni A Turunen | Helsinki University Hospital and University of Helsinki, Helsinki, Finland; Eye Genetics Group, Folkhälsan Research Center, Helsinki, Finland | | Clinical Groups | **Opthalmology Group** |
| Terhi Ollila | Hospital District of Helsinki and Uusimaa, Helsinki, Finland | | Clinical Groups | **Opthalmology Group** |
| Hannu Uusitalo | Pirkanmaa Hospital District, Tampere, Finland | | Clinical Groups | **Opthalmology Group** |
| Juha Karjalainen | Institute for Molecular Medicine Finland (FIMM), HiLIFE, University of Helsinki, Helsinki, Finland | | Clinical Groups | **Opthalmology Group** |
| Esa Pitkänen | Institute for Molecular Medicine Finland (FIMM), HiLIFE, University of Helsinki, Helsinki, Finland | | Clinical Groups | **Opthalmology Group** |
| Mengzhen Liu | Abbvie, Chicago, IL, United States | | Clinical Groups | **Opthalmology Group** |
| Heiko Runz | Biogen, Cambridge, MA, United States | | Clinical Groups | **Opthalmology Group** |
| Stephanie Loomis | Biogen, Cambridge, MA, United States | | Clinical Groups | **Opthalmology Group** |
| Erich Strauss | Genentech, San Francisco, CA, United States | | Clinical Groups | **Opthalmology Group** |
| Natalie Bowers | Genentech, San Francisco, CA, United States | | Clinical Groups | **Opthalmology Group** |
| Hao Chen | Genentech, San Francisco, CA, United States | | Clinical Groups | **Opthalmology Group** |
| Rion Pendergrass | Genentech, San Francisco, CA, United States | | Clinical Groups | **Opthalmology Group** |
| Kaisa Tasanen | Northern Ostrobothnia Hospital District, Oulu, Finland | | Clinical Groups | **Dermatology Group** |
| Laura Huilaja | Northern Ostrobothnia Hospital District, Oulu, Finland | | Clinical Groups | **Dermatology Group** |
| Katariina Hannula-Jouppi | Hospital District of Helsinki and Uusimaa, Helsinki, Finland | | Clinical Groups | **Dermatology Group** |
| Teea Salmi | Pirkanmaa Hospital District, Tampere, Finland | | Clinical Groups | **Dermatology Group** |
| Sirkku Peltonen | Hospital District of Southwest Finland, Turku, Finland | | Clinical Groups | **Dermatology Group** |
| Leena Koulu | Hospital District of Southwest Finland, Turku, Finland | | Clinical Groups | **Dermatology Group** |
| Nizar Smaoui | Abbvie, Chicago, IL, United States | | Clinical Groups | **Dermatology Group** |
| Fedik Rahimov | Abbvie, Chicago, IL, United States | | Clinical Groups | **Dermatology Group** |
| Anne Lehtonen | Abbvie, Chicago, IL, United States | | Clinical Groups | **Dermatology Group** |
| David Choy | Genentech, San Francisco, CA, United States | | Clinical Groups | **Dermatology Group** |
| Rion Pendergrass | Genentech, San Francisco, CA, United States | | Clinical Groups | **Dermatology Group** |
| Dawn Waterworth | Janssen Research & Development, LLC, Spring House, PA, United States | | Clinical Groups | **Dermatology Group** |
| Kirsi Kalpala | Pfizer, New York, NY, United States | | Clinical Groups | **Dermatology Group** |
| Ying Wu | Pfizer, New York, NY, United States | | Clinical Groups | **Dermatology Group** |
| Pirkko Pussinen | Hospital District of Helsinki and Uusimaa, Helsinki, Finland | | Clinical Groups | **Odontology Group** |
| Aino Salminen | Hospital District of Helsinki and Uusimaa, Helsinki, Finland | | Clinical Groups | **Odontology Group** |
| Tuula Salo | Hospital District of Helsinki and Uusimaa, Helsinki, Finland | | Clinical Groups | **Odontology Group** |
| David Rice | Hospital District of Helsinki and Uusimaa, Helsinki, Finland | | Clinical Groups | **Odontology Group** |
| Pekka Nieminen | Hospital District of Helsinki and Uusimaa, Helsinki, Finland | | Clinical Groups | **Odontology Group** |
| Ulla Palotie | Hospital District of Helsinki and Uusimaa, Helsinki, Finland | | Clinical Groups | **Odontology Group** |
| Maria Siponen | Northern Savo Hospital District, Kuopio, Finland | | Clinical Groups | **Odontology Group** |
| Liisa Suominen | Northern Savo Hospital District, Kuopio, Finland | | Clinical Groups | **Odontology Group** |
| Päivi Mäntylä | Northern Savo Hospital District, Kuopio, Finland | | Clinical Groups | **Odontology Group** |
| Ulvi Gursoy | Hospital District of Southwest Finland, Turku, Finland | | Clinical Groups | **Odontology Group** |
| Vuokko Anttonen | Northern Ostrobothnia Hospital District, Oulu, Finland | | Clinical Groups | **Odontology Group** |
| Kirsi Sipilä | Research Unit of Oral Health Sciences Faculty of Medicine, University of Oulu, Oulu, Finland; Medical Research Center, Oulu, Oulu University Hospital and University of Oulu, Oulu, Finland | | Clinical Groups | **Odontology Group** |
| Rion Pendergrass | Genentech, San Francisco, CA, United States | | Clinical Groups | **Odontology Group** |
| Hannele Laivuori | Institute for Molecular Medicine Finland (FIMM), HiLIFE, University of Helsinki, Helsinki, Finland | | Clinical Groups | **Women's Health and Reproduction Group** |
| Venla Kurra | Pirkanmaa Hospital District, Tampere, Finland | | Clinical Groups | **Women's Health and Reproduction Group** |
| Laura Kotaniemi-Talonen | Pirkanmaa Hospital District, Tampere, Finland | | Clinical Groups | **Women's Health and Reproduction Group** |
| Oskari Heikinheimo | Hospital District of Helsinki and Uusimaa, Helsinki, Finland | | Clinical Groups | **Women's Health and Reproduction Group** |
| Ilkka Kalliala | Hospital District of Helsinki and Uusimaa, Helsinki, Finland | | Clinical Groups | **Women's Health and Reproduction Group** |
| Lauri Aaltonen | Hospital District of Helsinki and Uusimaa, Helsinki, Finland | | Clinical Groups | **Women's Health and Reproduction Group** |
| Varpu Jokimaa | Hospital District of Southwest Finland, Turku, Finland | | Clinical Groups | **Women's Health and Reproduction Group** |
| Johannes Kettunen | Northern Ostrobothnia Hospital District, Oulu, Finland | | Clinical Groups | **Women's Health and Reproduction Group** |
| Marja Vääräsmäki | Northern Ostrobothnia Hospital District, Oulu, Finland | | Clinical Groups | **Women's Health and Reproduction Group** |
| Outi Uimari | Northern Ostrobothnia Hospital District, Oulu, Finland | | Clinical Groups | **Women's Health and Reproduction Group** |
| Laure Morin-Papunen | Northern Ostrobothnia Hospital District, Oulu, Finland | | Clinical Groups | **Women's Health and Reproduction Group** |
| Maarit Niinimäki | Northern Ostrobothnia Hospital District, Oulu, Finland | | Clinical Groups | **Women's Health and Reproduction Group** |
| Terhi Piltonen | Northern Ostrobothnia Hospital District, Oulu, Finland | | Clinical Groups | **Women's Health and Reproduction Group** |
| Katja Kivinen | Institute for Molecular Medicine Finland (FIMM), HiLIFE, University of Helsinki, Helsinki, Finland | | Clinical Groups | **Women's Health and Reproduction Group** |
| Elisabeth Widen | Institute for Molecular Medicine Finland (FIMM), HiLIFE, University of Helsinki, Helsinki, Finland | | Clinical Groups | **Women's Health and Reproduction Group** |
| Taru Tukiainen | Institute for Molecular Medicine Finland (FIMM), HiLIFE, University of Helsinki, Helsinki, Finland | | Clinical Groups | **Women's Health and Reproduction Group** |
| Mary Pat Reeve | Institute for Molecular Medicine Finland (FIMM), HiLIFE, University of Helsinki, Helsinki, Finland | | Clinical Groups | **Women's Health and Reproduction Group** |
| Mark Daly | Institute for Molecular Medicine Finland (FIMM), HiLIFE, University of Helsinki, Helsinki, Finland; Broad Institute of MIT and Harvard; Massachusetts General Hospital | | Clinical Groups | **Women's Health and Reproduction Group** |
| Niko Välimäki | University of Helsinki, Helsinki, Finland | | Clinical Groups | **Women's Health and Reproduction Group** |
| Eija Laakkonen | University of Jyväskylä, Jyväskylä, Finland | | Clinical Groups | **Women's Health and Reproduction Group** |
| Jaakko Tyrmi | University of Oulu, Oulu, Finland / University of Tampere, Tampere, Finland | | Clinical Groups | **Women's Health and Reproduction Group** |
| Heidi Silven | University of Oulu, Oulu, Finland | | Clinical Groups | **Women's Health and Reproduction Group** |
| Eeva Sliz | University of Oulu, Oulu, Finland | | Clinical Groups | **Women's Health and Reproduction Group** |
| Riikka Arffman | University of Oulu, Oulu, Finland | | Clinical Groups | **Women's Health and Reproduction Group** |
| Susanna Savukoski | University of Oulu, Oulu, Finland | | Clinical Groups | **Women's Health and Reproduction Group** |
| Triin Laisk | Estonian biobank, Tartu, Estonia | | Clinical Groups | **Women's Health and Reproduction Group** |
| Natalia Pujol | Estonian biobank, Tartu, Estonia | | Clinical Groups | **Women's Health and Reproduction Group** |
| Mengzhen Liu | Abbvie, Chicago, IL, United States | | Clinical Groups | **Women's Health and Reproduction Group** |
| Bridget Riley-Gillis | Abbvie, Chicago, IL, United States | | Clinical Groups | **Women's Health and Reproduction Group** |

| | | | | |
|---|---|---|---|---|
| Rion Pendergrass | Genentech, San Francisco, CA, United States | | Clinical Groups | **Women's Health and Reproduction Group** |
| Janet Kumar | GlaxoSmithKline, Collegeville, PA, United States | | Clinical Groups | **Women's Health and Reproduction Group** |
| Kirsi Auro | GlaxoSmithKline, Espoo, Finland | | Clinical Groups | **Women's Health and Reproduction Group** |
| Iiris Hovatta | University of Helsinki, Finland | | Clinical Groups | **Depression group** |
| Chia-Yen Chen | Biogen, Cambridge, MA, United States | | Clinical Groups | **Depression group** |
| Erkki Isometsä | Hospital District of Helsinki and Uusimaa, Helsinki, Finland | | Clinical Groups | **Depression group** |
| Hanna Ollila | Institute for Molecular Medicine Finland (FIMM), HiLIFE, University of Helsinki, Helsinki, Finland | | Clinical Groups | **Depression group** |
| Jaana Suvisaari | Finnish Institute for Health and Welfare (THL), Helsinki, Finland | | Clinical Groups | **Depression group** |
| Thomas Damm Als | Aarhus University, Denmark | | Clinical Groups | **Depression group** |
| Antti Mäkitie | Department of Otorhinolaryngology - Head and Neck Surgery, University of Helsinki and Helsinki University Hospital, Helsinki, Finland | | Clinical Groups | **ENT (ear, nose and throath) Group** |
| Argyro Bizaki-Vallaskangas | Pirkanmaa Hospital District, Tampere, Finland | | Clinical Groups | **ENT (ear, nose and throath) Group** |
| Sanna Toppila-Salmi | University of Helsinki, Finland | | Clinical Groups | **ENT (ear, nose and throath) Group** |
| Tytti Willberg | Hospital District of Southwest Finland, Turku, Finland | | Clinical Groups | **ENT (ear, nose and throath) Group** |
| Elmo Saarentaus | Institute for Molecular Medicine Finland (FIMM), HiLIFE, University of Helsinki, Helsinki, Finland | | Clinical Groups | **ENT (ear, nose and throath) Group** |
| Antti Aarnisalo | Hospital District of Helsinki and Uusimaa, Helsinki, Finland | | Clinical Groups | **ENT (ear, nose and throath) Group** |
| Eveliina Salminen | Hospital District of Helsinki and Uusimaa, Helsinki, Finland | | Clinical Groups | **ENT (ear, nose and throath) Group** |
| Elisa Rahikkala | Northern Ostrobothnia Hospital District, Oulu, Finland | | Clinical Groups | **ENT (ear, nose and throath) Group** |
| Johannes Kettunen | Northern Ostrobothnia Hospital District, Oulu, Finland | | Clinical Groups | **ENT (ear, nose and throath) Group** |
| Kristiina Aittomäki | Department of Medical Genetics, Helsinki University Central Hospital, Helsinki, Finland | | Clinical Groups | **POI (premature ovarian failure) Group** |
| Fredrik Åberg | Transplantation and Liver Surgery Clinic, Helsinki University Hospital, Helsinki University, Helsinki, Finland | | Clinical Groups | **LiverScore Group** |
| Mitja Kurki | Institute for Molecular Medicine Finland (FIMM), HiLIFE, University of Helsinki, Helsinki, Finland; Broad Institute, Cambridge, MA, United States | | **FinnGen Analysis working grou** | **FinnGen Analysis working group** |
| Samuli Ripatti | Institute for Molecular Medicine Finland (FIMM), HiLIFE, University of Helsinki, Helsinki, Finland | | **FinnGen Analysis working grou** | **FinnGen Analysis working group** |
| Mark Daly | Institute for Molecular Medicine, Finland (FIMM), HiLIFE, University of Helsinki, Helsinki, Finland; Broad Institute of MIT and Harvard; Massachusetts General Hospital | | **FinnGen Analysis working grou** | **FinnGen Analysis working group** |
| Juha Karjalainen | Institute for Molecular Medicine Finland (FIMM), HiLIFE, University of Helsinki, Helsinki, Finland | | **FinnGen Analysis working grou** | **FinnGen Analysis working group** |
| Aki Havulinna | Institute for Molecular Medicine Finland (FIMM), HiLIFE, University of Helsinki, Helsinki, Finland | | **FinnGen Analysis working grou** | **FinnGen Analysis working group** |
| Juha Mehtonen | Institute for Molecular Medicine Finland (FIMM), HiLIFE, University of Helsinki, Helsinki, Finland | | **FinnGen Analysis working grou** | **FinnGen Analysis working group** |
| Priit Palta | Institute for Molecular Medicine Finland (FIMM), HiLIFE, University of Helsinki, Helsinki, Finland | | **FinnGen Analysis working grou** | **FinnGen Analysis working group** |
| Shabbeer Hassan | Institute for Molecular Medicine Finland (FIMM), HiLIFE, University of Helsinki, Helsinki, Finland | | **FinnGen Analysis working grou** | **FinnGen Analysis working group** |
| Pietro Della Briotta Parolo | Institute for Molecular Medicine Finland (FIMM), HiLIFE, University of Helsinki, Helsinki, Finland | | **FinnGen Analysis working grou** | **FinnGen Analysis working group** |
| Wei Zhou | Broad Institute, Cambridge, MA, United States | | **FinnGen Analysis working grou** | **FinnGen Analysis working group** |
| Mutaamba Maasha | Broad Institute, Cambridge, MA, United States | | **FinnGen Analysis working grou** | **FinnGen Analysis working group** |
| Shabbeer Hassan | Institute for Molecular Medicine Finland (FIMM), HiLIFE, University of Helsinki, Helsinki, Finland | | **FinnGen Analysis working grou** | **FinnGen Analysis working group** |
| Susanna Lemmelä | Institute for Molecular Medicine Finland (FIMM), HiLIFE, University of Helsinki, Helsinki, Finland | | **FinnGen Analysis working grou** | **FinnGen Analysis working group** |
| Manuel Rivas | University of Stanford, Stanford, CA, United States | | **FinnGen Analysis working grou** | **FinnGen Analysis working group** |
| Aarno Palotie | Institute for Molecular Medicine Finland (FIMM), HiLIFE, University of Helsinki, Helsinki, Finland | | **FinnGen Analysis working grou** | **FinnGen Analysis working group** |
| Aoxing Liu | Institute for Molecular Medicine Finland (FIMM), HiLIFE, University of Helsinki, Helsinki, Finland | | **FinnGen Analysis working grou** | **FinnGen Analysis working group** |
| Arto Lehisto | Institute for Molecular Medicine Finland (FIMM), HiLIFE, University of Helsinki, Helsinki, Finland | | **FinnGen Analysis working grou** | **FinnGen Analysis working group** |
| Andrea Ganna | Institute for Molecular Medicine Finland (FIMM), HiLIFE, University of Helsinki, Helsinki, Finland | | **FinnGen Analysis working grou** | **FinnGen Analysis working group** |
| Vincent Llorens | Institute for Molecular Medicine Finland (FIMM), HiLIFE, University of Helsinki, Helsinki, Finland | | **FinnGen Analysis working grou** | **FinnGen Analysis working group** |
| Hannele Laivuori | Institute for Molecular Medicine Finland (FIMM), HiLIFE, University of Helsinki, Helsinki, Finland | | **FinnGen Analysis working grou** | **FinnGen Analysis working group** |
| Taru Tukiainen | Institute for Molecular Medicine Finland (FIMM), HiLIFE, University of Helsinki, Helsinki, Finland | | **FinnGen Analysis working grou** | **FinnGen Analysis working group** |
| Mary Pat Reeve | Institute for Molecular Medicine Finland (FIMM), HiLIFE, University of Helsinki, Helsinki, Finland | | **FinnGen Analysis working grou** | **FinnGen Analysis working group** |
| Henrike Heyne | Institute for Molecular Medicine Finland (FIMM), HiLIFE, University of Helsinki, Helsinki, Finland | | **FinnGen Analysis working grou** | **FinnGen Analysis working group** |
| Nina Mars | Institute for Molecular Medicine Finland (FIMM), HiLIFE, University of Helsinki, Helsinki, Finland | | **FinnGen Analysis working grou** | **FinnGen Analysis working group** |
| Joel Rämö | Institute for Molecular Medicine Finland (FIMM), HiLIFE, University of Helsinki, Helsinki, Finland | | **FinnGen Analysis working grou** | **FinnGen Analysis working group** |
| Elmo Saarentaus | Institute for Molecular Medicine Finland (FIMM), HiLIFE, University of Helsinki, Helsinki, Finland | | **FinnGen Analysis working grou** | **FinnGen Analysis working group** |
| Hanna Ollila | Institute for Molecular Medicine Finland (FIMM), HiLIFE, University of Helsinki, Helsinki, Finland | | **FinnGen Analysis working grou** | **FinnGen Analysis working group** |
| Rodos Rodosthenous | Institute for Molecular Medicine Finland (FIMM), HiLIFE, University of Helsinki, Helsinki, Finland | | **FinnGen Analysis working grou** | **FinnGen Analysis working group** |
| Satu Strausz | Institute for Molecular Medicine Finland (FIMM), HiLIFE, University of Helsinki, Helsinki, Finland | | **FinnGen Analysis working grou** | **FinnGen Analysis working group** |
| Tuula Palotie | University of Helsinki and Hospital District of Helsinki and Uusimaa, Helsinki, Finland | | **FinnGen Analysis working grou** | **FinnGen Analysis working group** |
| Kimmo Palin | University of Helsinki, Helsinki, Finland | | **FinnGen Analysis working grou** | **FinnGen Analysis working group** |
| Javier Garcia-Tabuenca | University of Tampere, Tampere, Finland | | **FinnGen Analysis working grou** | **FinnGen Analysis working group** |
| Harri Siirtola | University of Tampere, Tampere, Finland | | **FinnGen Analysis working grou** | **FinnGen Analysis working group** |
| Tuomo Kiiskinen | Institute for Molecular Medicine Finland (FIMM), HiLIFE, University of Helsinki, Helsinki, Finland | | **FinnGen Analysis working grou** | **FinnGen Analysis working group** |
| Jiwoo Lee | Institute for Molecular Medicine Finland (FIMM), HiLIFE, University of Helsinki, Helsinki, Finland; Broad Institute, Cambridge, MA, United States | | **FinnGen Analysis working grou** | **FinnGen Analysis working group** |
| Kristin Tsuo | Institute for Molecular Medicine Finland (FIMM), HiLIFE, University of Helsinki, Helsinki, Finland; Broad Institute, Cambridge, MA, United States | | **FinnGen Analysis working grou** | **FinnGen Analysis working group** |
| Amanda Elliott | Institute for Molecular Medicine Finland (FIMM), HiLIFE, University of Helsinki, Helsinki, Finland; Broad Institute, Cambridge, MA, USA and Massachusetts General Hospital, Boston, MA, USA | | **FinnGen Analysis working grou** | **FinnGen Analysis working group** |
| Kati Kristiansson | THL Biobank / Finnish Institute for Health and Welfare (THL), Helsinki, Finland | | **FinnGen Analysis working grou** | **FinnGen Analysis working group** |
| Mikko Arvas | Finnish Red Cross Blood Service / Finnish Hematology Registry and Clinical Biobank, Helsinki, Finland | | **FinnGen Analysis working grou** | **FinnGen Analysis working group** |
| Kati Hyvärinen | Finnish Red Cross Blood Service, Helsinki, Finland | | **FinnGen Analysis working grou** | **FinnGen Analysis working group** |
| Jarmo Ritari | Finnish Red Cross Blood Service, Helsinki, Finland | | **FinnGen Analysis working grou** | **FinnGen Analysis working group** |
| Olli Carpén | Helsinki Biobank / Helsinki University and Hospital District of Helsinki and Uusimaa, Helsinki | | **FinnGen Analysis working grou** | **FinnGen Analysis working group** |
| Johannes Kettunen | Northern Finland Biobank Borealis / University of Oulu / Northern Ostrobothnia Hospital District, Oulu, Finland | | **FinnGen Analysis working grou** | **FinnGen Analysis working group** |
| Katri Pylkäs | University of Oulu, Oulu, Finland | | **FinnGen Analysis working grou** | **FinnGen Analysis working group** |
| Eeva Sliz | University of Oulu, Oulu, Finland | | **FinnGen Analysis working grou** | **FinnGen Analysis working group** |
| Minna Karjalainen | University of Oulu, Oulu, Finland | | **FinnGen Analysis working grou** | **FinnGen Analysis working group** |
| Tuomo Mantere | Northern Finland Biobank Borealis / University of Oulu / Northern Ostrobothnia Hospital District, Oulu, Finland | | **FinnGen Analysis working grou** | **FinnGen Analysis working group** |
| Eeva Kangasniemi | Finnish Clinical Biobank Tampere / University of Tampere / Pirkanmaa Hospital District, Tampere, Finland | | **FinnGen Analysis working grou** | **FinnGen Analysis working group** |
| Sami Heikkinen | University of Eastern Finland, Kuopio, Finland | | **FinnGen Analysis working grou** | **FinnGen Analysis working group** |
| Arto Mannermaa | Biobank of Eastern Finland / University of Eastern Finland / Northern Savo Hospital District, Kuopio, Finland | | **FinnGen Analysis working grou** | **FinnGen Analysis working group** |
| Eija Laakkonen | University of Jyväskylä, Jyväskylä, Finland | | **FinnGen Analysis working grou** | **FinnGen Analysis working group** |
| Nina Pitkänen | Auria Biobank / University of Turku / Hospital District of Southwest Finland, Turku, Finland | | **FinnGen Analysis working grou** | **FinnGen Analysis working group** |
| Samuel Lessard | Translational Sciences, Sanofi R&D, Framingham, MA, USA | | **FinnGen Analysis working grou** | **FinnGen Analysis working group** |
| Clément Chatelain | Translational Sciences, Sanofi R&D, Framingham, MA, USA | | **FinnGen Analysis working grou** | **FinnGen Analysis working group** |
| Lila Kallio | Auria Biobank / University of Turku / Hospital District of Southwest Finland, Turku, Finland | | **Biobank directors** | **Biobank directors** |
| Tiina Wahlfors | THL Biobank / Finnish Institute for Health and Welfare (THL), Helsinki, Finland | | **Biobank directors** | **Biobank directors** |
| Jukka Partanen | Finnish Red Cross Blood Service / Finnish Hematology Registry and Clinical Biobank, Helsinki, Finland | | **Biobank directors** | **Biobank directors** |
| Eero Punkka | Helsinki Biobank / Helsinki University and Hospital District of Helsinki and Uusimaa, Helsinki | | **Biobank directors** | **Biobank directors** |
| Raisa Serpi | Northern Finland Biobank Borealis / University of Oulu / Northern Ostrobothnia Hospital District, Oulu, Finland | | **Biobank directors** | **Biobank directors** |
| Sanna Siltanen | Finnish Clinical Biobank Tampere / University of Tampere / Pirkanmaa Hospital District, Tampere, Finland | | **Biobank directors** | **Biobank directors** |
| Veli-Matti Kosma | Biobank of Eastern Finland / University of Eastern Finland / Northern Savo Hospital District, Kuopio, Finland | | **Biobank directors** | **Biobank directors** |
| Teijo Kuopio | Central Finland Biobank / University of Jyväskylä / Central Finland Health Care District, Jyväskylä, Finland | | **Biobank directors** | **Biobank directors** |
| Anu Jalanko | Institute for Molecular Medicine Finland (FIMM), HiLIFE, University of Helsinki, Helsinki, Finland | | **FinnGen Teams** | **Administration** |
| Huei-Yi Shen | Institute for Molecular Medicine Finland (FIMM), HiLIFE, University of Helsinki, Helsinki, Finland | | **FinnGen Teams** | **Administration** |
| Risto Kajanne | Institute for Molecular Medicine Finland (FIMM), HiLIFE, University of Helsinki, Helsinki, Finland | | **FinnGen Teams** | **Administration** |
| Mervi Aavikko | Institute for Molecular Medicine Finland (FIMM), HiLIFE, University of Helsinki, Helsinki, Finland | | **FinnGen Teams** | **Administration** |
| Rasko Leinonen | Institute for Molecular Medicine Finland (FIMM), HiLIFE, University of Helsinki, Helsinki, Finland; | | **FinnGen Teams** | **Administration** |
| Henna Palin | Finnish Clinical Biobank Tampere / University of Tampere / Pirkanmaa Hospital District, Tampere, Finland | | **FinnGen Teams** | **Administration** |
| Malla-Maria Linna | Helsinki Biobank / Helsinki University and Hospital District of Helsinki and Uusimaa, Helsinki | | **FinnGen Teams** | **Administration** |
| Mitja Kurki | Institute for Molecular Medicine Finland (FIMM), HiLIFE, University of Helsinki, Helsinki, Finland; Broad Institute, Cambridge, MA, United States | | **FinnGen Teams** | **Analysis** |
| Juha Karjalainen | Institute for Molecular Medicine Finland (FIMM), HiLIFE, University of Helsinki, Helsinki, Finland | | **FinnGen Teams** | **Analysis** |

| | | | | |
|---|---|---|---|---|
| Pietro Della Briotta Parolo | Institute for Molecular Medicine Finland (FIMM), HiLIFE, University of Helsinki, Helsinki, Finland | | **FinnGen Teams** | **Analysis** |
| Arto Lehisto | Institute for Molecular Medicine Finland (FIMM), HiLIFE, University of Helsinki, Helsinki, Finland | | **FinnGen Teams** | **Analysis** |
| Juha Mehtonen | Institute for Molecular Medicine Finland (FIMM), HiLIFE, University of Helsinki, Helsinki, Finland | | **FinnGen Teams** | **Analysis** |
| Wei Zhou | Broad Institute, Cambridge, MA, United States | | **FinnGen Teams** | **Analysis** |
| Masahiro Kanai | Broad Institute, Cambridge, MA, United States | | **FinnGen Teams** | **Analysis** |
| Mutaamba Maasha | Broad Institute, Cambridge, MA, United States | | **FinnGen Teams** | **Analysis** |
| Zhili Zheng | Broad Institute, Cambridge, MA, United States | | **FinnGen Teams** | **Analysis** |
| Hannele Laivuori | Institute for Molecular Medicine Finland (FIMM), HiLIFE, University of Helsinki, Helsinki, Finland | | **FinnGen Teams** | **Clinical Endpoint Development** |
| Aki Havulinna | Institute for Molecular Medicine Finland (FIMM), HiLIFE, University of Helsinki, Helsinki, Finland | | **FinnGen Teams** | **Clinical Endpoint Development** |
| Susanna Lemmelä | Institute for Molecular Medicine Finland (FIMM), HiLIFE, University of Helsinki, Helsinki, Finland | | **FinnGen Teams** | **Clinical Endpoint Development** |
| Tuomo Kiiskinen | Institute for Molecular Medicine Finland (FIMM), HiLIFE, University of Helsinki, Helsinki, Finland | | **FinnGen Teams** | **Clinical Endpoint Development** |
| L. Elisa Lahtela | Institute for Molecular Medicine Finland (FIMM), HiLIFE, University of Helsinki, Helsinki, Finland | | **FinnGen Teams** | **Clinical Endpoint Development** |
| Mari Kaunisto | Institute for Molecular Medicine Finland (FIMM), HiLIFE, University of Helsinki, Helsinki, Finland | | **FinnGen Teams** | **Communication** |
| Elina Kilpeläinen | Institute for Molecular Medicine Finland (FIMM), HiLIFE, University of Helsinki, Helsinki, Finland | | **FinnGen Teams** | **E-Science** |
| Timo P. Sipilä | Institute for Molecular Medicine Finland (FIMM), HiLIFE, University of Helsinki, Helsinki, Finland | | **FinnGen Teams** | **E-Science** |
| Oluwaseun Alexander Dada | Institute for Molecular Medicine Finland (FIMM), HiLIFE, University of Helsinki, Helsinki, Finland | | **FinnGen Teams** | **E-Science** |
| Awaisa Ghazal | Institute for Molecular Medicine Finland (FIMM), HiLIFE, University of Helsinki, Helsinki, Finland | | **FinnGen Teams** | **E-Science** |
| Anastasia Kytölä | Institute for Molecular Medicine Finland (FIMM), HiLIFE, University of Helsinki, Helsinki, Finland | | **FinnGen Teams** | **E-Science** |
| Rigbe Weldatsadik | Institute for Molecular Medicine Finland (FIMM), HiLIFE, University of Helsinki, Helsinki, Finland | | **FinnGen Teams** | **E-Science** |
| Sanni Ruotsalainen | Institute for Molecular Medicine Finland (FIMM), HiLIFE, University of Helsinki, Helsinki, Finland | | **FinnGen Teams** | **E-Science** |
| Kati Donner | Institute for Molecular Medicine Finland (FIMM), HiLIFE, University of Helsinki, Helsinki, Finland | | **FinnGen Teams** | **Genotyping** |
| Timo P. Sipilä | Institute for Molecular Medicine Finland (FIMM), HiLIFE, University of Helsinki, Helsinki, Finland | | **FinnGen Teams** | **Genotyping** |
| Anu Loukola | Helsinki Biobank / Helsinki University and Hospital District of Helsinki and Uusimaa, Helsinki | | **FinnGen Teams** | **Sample Collection Coordination** |
| Päivi Laiho | THL Biobank / Finnish Institute for Health and Welfare (THL), Helsinki, Finland | | **FinnGen Teams** | **Sample Logistics** |
| Tuuli Sistonen | THL Biobank / Finnish Institute for Health and Welfare (THL), Helsinki, Finland | | **FinnGen Teams** | **Sample Logistics** |
| Essi Kaiharju | THL Biobank / Finnish Institute for Health and Welfare (THL), Helsinki, Finland | | **FinnGen Teams** | **Sample Logistics** |
| Markku Laukkanen | THL Biobank / Finnish Institute for Health and Welfare (THL), Helsinki, Finland | | **FinnGen Teams** | **Sample Logistics** |
| Elina Järvensivu | THL Biobank / Finnish Institute for Health and Welfare (THL), Helsinki, Finland | | **FinnGen Teams** | **Sample Logistics** |
| Sini Lähteenmäki | THL Biobank / Finnish Institute for Health and Welfare (THL), Helsinki, Finland | | **FinnGen Teams** | **Sample Logistics** |
| Lotta Männikkö | THL Biobank / Finnish Institute for Health and Welfare (THL), Helsinki, Finland | | **FinnGen Teams** | **Sample Logistics** |
| Regis Wong | THL Biobank / Finnish Institute for Health and Welfare (THL), Helsinki, Finland | | **FinnGen Teams** | **Sample Logistics** |
| Auli Toivola | THL Biobank / Finnish Institute for Health and Welfare (THL), Helsinki, Finland | | **FinnGen Teams** | **Sample Logistics** |
| Minna Brunfeldt | THL Biobank / Finnish Institute for Health and Welfare (THL), Helsinki, Finland | | **FinnGen Teams** | **Registry Data Operations** |
| Hannele Mattsson | THL Biobank / Finnish Institute for Health and Welfare (THL), Helsinki, Finland | | **FinnGen Teams** | **Registry Data Operations** |
| Kati Kristiansson | THL Biobank / Finnish Institute for Health and Welfare (THL), Helsinki, Finland | | **FinnGen Teams** | **Registry Data Operations** |
| Susanna Lemmelä | Institute for Molecular Medicine Finland (FIMM), HiLIFE, University of Helsinki, Helsinki, Finland | | **FinnGen Teams** | **Registry Data Operations** |
| Sami Koskelainen | THL Biobank / Finnish Institute for Health and Welfare (THL), Helsinki, Finland | | **FinnGen Teams** | **Registry Data Operations** |
| Tero Hiekkalinna | THL Biobank / Finnish Institute for Health and Welfare (THL), Helsinki, Finland | | **FinnGen Teams** | **Registry Data Operations** |
| Teemu Paajanen | THL Biobank / Finnish Institute for Health and Welfare (THL), Helsinki, Finland | | **FinnGen Teams** | **Registry Data Operations** |
| Priit Palta | Institute for Molecular Medicine Finland (FIMM), HiLIFE, University of Helsinki, Helsinki, Finland | | **FinnGen Teams** | **Sequencing Informatics** |
| Kalle Pärn | Institute for Molecular Medicine Finland (FIMM), HiLIFE, University of Helsinki, Helsinki, Finland | | **FinnGen Teams** | **Sequencing Informatics** |
| Mart Kals | Institute for Molecular Medicine Finland (FIMM), HiLIFE, University of Helsinki, Helsinki, Finland | | **FinnGen Teams** | **Sequencing Informatics** |
| Shuang Luo | Institute for Molecular Medicine Finland (FIMM), HiLIFE, University of Helsinki, Helsinki, Finland | | **FinnGen Teams** | **Sequencing Informatics** |
| Tarja Laitinen | Pirkanmaa Hospital District, Tampere, Finland | | **FinnGen Teams** | **Trajectory** |
| Mary Pat Reeve | Institute for Molecular Medicine Finland (FIMM), HiLIFE, University of Helsinki, Helsinki, Finland | | **FinnGen Teams** | **Trajectory** |
| Shanmukha Sampath Padm | Institute for Molecular Medicine Finland (FIMM), HiLIFE, University of Helsinki, Helsinki, Finland | | **FinnGen Teams** | **Trajectory** |
| Marianna Niemi | University of Tampere, Tampere, Finland | | **FinnGen Teams** | **Trajectory** |
| Harri Siirtola | University of Tampere, Tampere, Finland | | **FinnGen Teams** | **Trajectory** |
| Javier Gracia-Tabuenca | University of Tampere, Tampere, Finland | | **FinnGen Teams** | **Trajectory** |
| Mika Helminen | University of Tampere, Tampere, Finland | | **FinnGen Teams** | **Trajectory** |
| Tiina Luukkaala | University of Tampere, Tampere, Finland | | **FinnGen Teams** | **Trajectory** |
| Iida Vähätalo | University of Tampere, Tampere, Finland | | **FinnGen Teams** | **Trajectory** |
| Jyrki Tammerluoto | Institute for Molecular Medicine Finland (FIMM), HiLIFE, University of Helsinki, Helsinki, Finland | | **FinnGen Teams** | **Data protection officer** |
| Marco Hautalahti | Finnish Biobank Cooperative - FINBB | | **FinnGen Teams** | **FINBB - Finnish biobank cooperative** |
| Johanna Mäkelä | Finnish Biobank Cooperative - FINBB | | **FinnGen Teams** | **FINBB - Finnish biobank cooperative** |
| Sarah Smith | Finnish Biobank Cooperative - FINBB | | **FinnGen Teams** | **FINBB - Finnish biobank cooperative** |
| Tom Southerington | Finnish Biobank Cooperative - FINBB | | **FinnGen Teams** | **FINBB - Finnish biobank cooperative** |
| Petri Lehto | Finnish Biobank Cooperative - FINBB | | **FinnGen Teams** | **FINBB - Finnish biobank cooperative** |
